# Rising rates of young people Not in Education, Employment, or Training (NEET) explained by higher prevalence of physical and psychological ill health: a 15-year UK study

**DOI:** 10.64898/2026.08.11.26360217

**Authors:** Jacques Wels, Dominic Kelly, Demelza Smeeth, Charis Bridger-Staatz, Zixu Li, George Ploubidis, Nishi Chaturvedi, Praveetha Patalay

## Abstract

**Background:** Rising rates of young people Not in Education, Employment, or Training (NEET) in the UK have recently coincided with declining youth physical and mental health but no study has asked whether this reflects a growing proportion of young people with health problems (prevalence) or those with health problems becoming more likely to be NEET (penalty).

**Methods:** Using 15 years of Understanding Society data (2009-23), we analysed 15,242 respondents aged 16-24 (66,160 observations). We employed three complementary approaches: descriptive trends, Blinder-Oaxaca-Kitagawa (BO) probit decomposition comparing 2009-2013 and 2019-2023 against a 2014-2018 reference period, and fixed-effects (FE) Poisson models with lagged health status. Exposures included self-reported health conditions or disability (SRHD), psychological distress, diagnosed conditions and socio- demographic factors.

**Findings:** NEET rates were lowest in 2014-18 (10.5-11.5%) and higher in 2009-13 (12-15%) and 2019-23 (15-16%). Higher prevalence of SRHD, psychological distress, diagnosed depression and multimorbidity explained changes in NEET prevalence across both the 2009- 13 to 2014-2018 and 2014-18 to 2019-23 periods. No change in penalty was observed for any health variable across periods, except for an increase in the penalty for SRHD between the 2009-13 to 2014-18 periods.

**Interpretation:** Rising NEET rates among UK youth are driven largely by more young people having physical and psychological ill health. Whilst labour market and education accommodations remain important, reducing NEET rates will require reversing the decline in youth health, not just accommodating it.

## Background

Young people in the United Kingdom face a dual challenge: increasing numbers Not in Employment, Education, or Training (NEET) ^1^ and declining health, both mental and physical. ^2^ However, no study has asked whether the link from poor health to NEET reflects a growing proportion of youth with ill health, or whether those with ill health are now more likely to become NEET. This distinction matters because the two mechanisms imply different interventions: one targets the health crisis itself, the other targets the labour market barriers faced by young people with poor mental health.

In the UK, it is estimated that around 13% of all 16-24 year olds are currently NEET.^1,3^ This is particularly concerning as being out of employment is linked to a whole host of social, economic and health adversities, creating a vicious cycle.^4^ In response to these challenges, the UK government has unveiled a “Youth Guarantee” plan for 18-21 year olds, promising access to education, training, or help to find a job. ^5^ This initiative, part of the “Get Britain Working” policy, explicitly aims to address multiple drivers, with a special focus on worsening mental health.^6^

Most studies on the NEET phenomenon have leveraged longitudinal data to address the influence of earlier health and other predictors of later NEET status. Socio-economic factors such as growing up in an unstable home environment or having parents with low educational attainment or who are workless ^7^ as well as residing in deprived neighbourhoods ^8^ are associated with increased risks of becoming NEET.^9^ Similarly, childcare responsibilities, disproportionately affect women.^10^ One factor contributing to the long-term reduction in female NEETs has been fewer women having children at younger ages ^11^ but obstacles for young mothers persist.^12^ Looking at health; congenital conditions and diseases acquired from childhood into adulthood are significant barriers to educational and labour market participation ^13^. Psychological factors such as a lack of a robust sense of self, limited self-efficacy, and risky behaviours are also prominent risk factors.^9^

The number of young NEET people has fallen since the 1970s but there has been an increase around 2008-2013 linked to the financial crash and recession, then a period of lower rates from 2014 to 2018 and finally growing rates over recent years.^1,11^ Understanding this recent increase requires careful attention to two distinct sources of change: shifts in the composition of the young population across different characteristics, critically the prevalence of mental health issues, and changes in the strength of the association between those characteristics and the risk of becoming NEET.

The recent increase in NEET rates is recognised to be partially driven by the declining health of young people ^1^ but little is known on whether this effect is due to the increase in health conditions or changes in the consequences of health conditions associated with greater risks of being NEET. This study aimed to determine whether the recent growing NEET levels are due to increasing prevalence of physical and mental health conditions, and/or the strength of the association between health conditions and becoming a NEET. Using a combination of methods on a large UK representative dataset covering years 2009 to 2023, we ask (1) whether greater prevalence in physical and mental ill health explains increasing NEET rates; (2) whether the penalty associated with ill health has changed between 2009 and 2023 and explains increasing NEET rates; and (3) what are the longitudinal associations between individual NEET risks and ill health and have these associations changed over time?

## Data and methods

### Data

We used data from the UK Household Longitudinal Study (UKHLS – Understanding Society) across waves 2009–10 to 2022-23, ^14^ restricting the sample to respondents aged 16– 24 at each wave. This nationally representative survey provides robust, repeated measures of NEET status and physical and mental health, and offers the longest continuous observation window available for this age group.

### Outcome

The primary outcome is NEET status, defined as not being in education, employment, or training. NEET was derived from the labour force status variable and coded as a binary indicator where 1 represents NEET and 0 represents all other states. Individuals were classified as NEET if they were unemployed, long-term sick or disabled, or reported “doing something else”. The reference category of non-NEET includes all other statuses: employed (including self-employment and furlough ^15^); in education, training or apprenticeship and other (maternity leave, family care, unpaid family workers, parental or adoption leave).

### Exposures

The study includes two blocks of exposure variables, one related to health and the other related demographic and socio-economic factors.

Health-related variables include self-reported health condition or disability (yes or no) and psychological distress caseness assessed using the 12-item General Health Questionnaire (GHQ-12) where scores of 4 or more were classified as indicating psychological distress.^16^ The survey additionally includes information on specific ever diagnosed health conditions and whether new conditions were diagnosed over the survey period. This information was not asked to respondents entering the survey in 2010-11, reducing the sample size, as can be seen in <u>supplementary file S1.</u> However, it does not compromise the representativeness of the sample as no significant association was observed between the selection criteria and exposure factors (<u>supplementary file S2</u>). We provide results for the following conditions: asthma, depression, diabetes, epilepsy and high blood pressure. We additionally generated a multimorbidity variable distinguishing respondents with two or more health conditions versus one or none, calculated on the above-mentioned conditions plus chronic bronchitis, hypothyroidism, cancer or malignancy and liver condition for which we do not provide individual estimates because of low counts.

Demographic and socio-economic factors include age (16-17 vs. 18-24, ref.), sex (ref.: male), ethnicity (white vs. non-white, ref.), highest level of education (GCSE or equivalent vs. a qualification above GCSE, ref.), the region of residence (London and the South of England vs. the rest of the country, ref.), whether the respondent has the responsibility of a child aged less than 14 (yes vs. no, ref.). We also include the equivalized household incomes quintiles to adjust household incomes based on household composition. We applied the modified OECD equivalence scale, which assigns a value of 1 to the first adult, 0.5 to each subsequent adult, and 0.3 to each child. ^17^ Equivalised income was then calculated as total household net income divided by the equivalence factor. These quintiles were calculated on a yearly basis to account for inflation. The lowest quintile represents the poorest 20 percent. Finally, the Index of multiple deprivation (IMD) ^18^ was measured separately across UK countries and represents a combined measure (in quintiles) of deprivation across seven domains: income, employment, education, health, crime, access to services and housing environment.

## Methods

We employed three complementary approaches to examine the health-NEET relationship across periods.

First, we examined trends in NEET probabilities over time using modified Poisson regression with robust standard errors. ^19,20^ This approach directly estimates risk ratios rather than odds ratios, which is advantageous when the outcome is common. For each wave from 2009 to 2023, we estimated predicted cross-sectional probabilities of being NEET for each exposure and additionally provided yearly prevalence and 95% confidence interval (95%CI) of each exposure.

Second, to investigate the sources of differences in NEET rates between periods, we employed univariate Blinder-Oaxaca-Kitagawa (BO) ^21–24^ decomposition to decompose outcome gaps between groups into components attributable to differences in characteristics and differences in the effects of those characteristics. The decomposition begins by estimating separate probit regression models for each period, modelling the probability of being NEET as a function of the explanatory variables. The difference in mean NEET rates between two periods is then partitioned into two components. The *explained component* captures the part of the gap that would be expected given differences in the observed characteristics between the two periods. The *unexplained component* captures the part of the gap due to differences in the coefficients (or penalty) of those characteristics.

We conducted two separate decompositions. The first decomposition compares periods 2009- 2013 and 2019-2023 to 2014-2018 (where NEET levels were at their lowest). The second decomposition, used for visualisation, compares each year to 2016 (i.e., the middle year). The decomposition coefficients are reported as percentage points (probability-scale coefficients × 100).

Third, we estimate univariate Fixed Effects Modified Poisson (FE) models with a multiplicative interaction between one-wave lagged exposure and the three periods (2009- 2013, 2019-2023, 2014-2018 (ref.)), allowing the exposure effect to differ across time. As for the decomposition, we replicated the model with a year interaction using 2016 as the reference category. The fixed effects remove all time-invariant unobserved differences between individuals which provides the within-person effect of lagged exposures on NEET for each period. The analytical sample is restricted to respondents with at least two consecutive waves. This provides an estimate comparable to BO’s unexplained component. If both methods point in the same direction, we can be more confident that BO’s unexplained component reflects genuine period-specific processes rather than omitted stable characteristics or state dependence. Fixed-effects The estimates are reported as raw coefficients, which range from -∞ to +∞ and are therefore directly comparable to the logit BO coefficients.

All models were run separately with a single exposure to avoid overadjustment. ^25^ We used cross-sectional weights in BO and robust variance estimation to account for clustered data in all models. The R software (version 4.2.3) was used data cleaning, descriptive statistics and plots. Stata (version 18.5) was used for decomposition and fixed-effects modelling.

### Additional analyses

To address potential overlap between self-reported health condition or disability and psychological distress, we replicated the decomposition and fixed effects models conjointly adjusting for both variables. To address sub-group differences in NEET prevalence and penalty, we stratified the BO decomposition using 2016 as the reference category across socio-demographic factors. We additionally replicated the FE for sex and age-group (16-19 and 20-24).

## Results

The total sample includes 66,160 observations (15,242 respondents) with different levels of missing values for health conditions or disability. For instance, 56,242 (14,429 respondents) had information on GHQ-12. Due to specific health conditions variables not collected at survey entry on wave 2, the sample has 49,932 observations (12,147 respondents). Lagged models further restricted the sample to 70 percent of the original samples and 94 percent of the analytic samples, as can be seen in <u>supplementary file S1</u>.

### Prevalence and risk of being NEET

**Figure 1** shows the annual percentage of young people aged 16–24 years classified as NEET alongside the composition of both the NEET and non-NEET subgroups from 2009 to 2023. Three distinct trends emerge. During the mid-period (2014–2018), the probability of being NEET was relatively low, affecting 10.5–11.5% of this population – a time when the age of compulsory education or training in England increased to 17 and then to 18 years. During the Great Recession (2009–2013), NEET prevalence ranged from 15% in 2009 to 12% in 2013. In the COVID-19 and post-COVID-19 period, NEET proportions reached 15% in 2020 and 16% in 2023.

**Figure 1.**
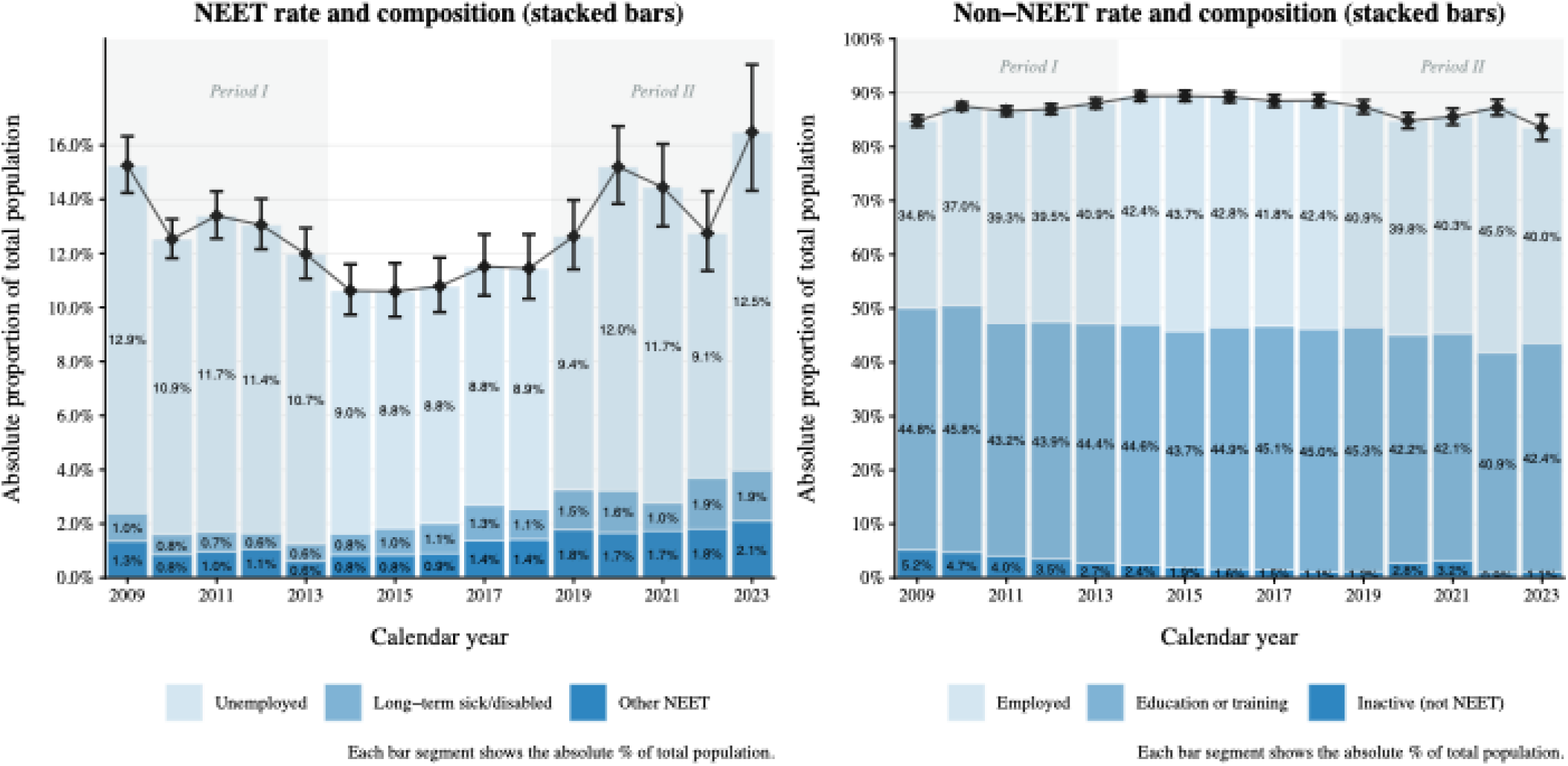
NEET and non-NEET rates and Composition of the 16-24 population per year (2009-2023)

Among NEETs, the percentage of respondents reporting unemployment began to decline in 2014 and increased again in 2019 reaching 12.5 percent of the 16-24 population in 2023. By contrast, the proportion reporting long-term sickness or disability almost doubled over the study period, from 1.0 percent in 2009 to 1.9 percent in 2023. A similar increase was seen for those reporting doing something else (other NEET). Within the non-NEET population, the percentage of respondents in education or training fell over time, whereas the share of those in employment (employed and self-employed) within the non-NEET category increased by about 5 percentage points between 2009 and 2023.

**Figure 2** exhibits the factors’ prevalences and risk ratios by year. One observes a steady increase in the prevalence of health condition or disability and psychological distress starting around 2017, which has amplified since then, while the corresponding relative risks have slightly increased over the same time period. Those with two or more conditions are a small proportion within the 16-24 populations and the prevalence of multimorbidity has declined until 2019, mainly driven by a reduction in asthma, and increased since 2020, mainly driven by clinical depression. However, the relative risks of being NEET among those with multimorbidity remained stable. Among the socio-demographic factors shown in <u>supplementary file S3</u>, the risk of being NEET for those aged 16-17 compared to those aged 18-24 has constantly increased since 2016, although is still smaller than for the 18-24 age group. The proportion with childcare responsibilities has declined over time, but such responsibilities continue to be associated with a non-significant increase in risk.

**Figure 2.**
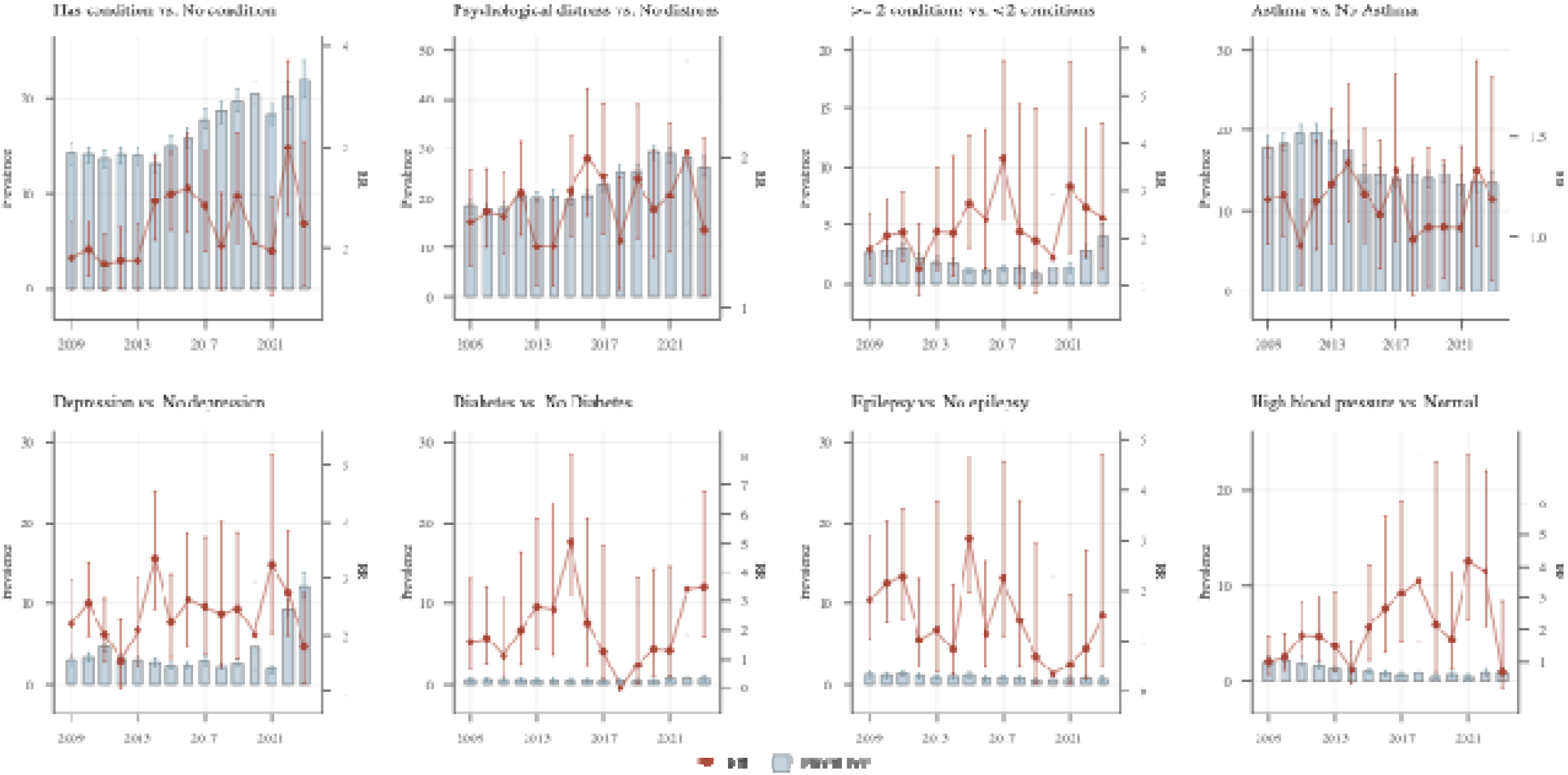
Prevalence and Relative Risks (RR) of being NEET by health conditions between 2009 and 2023.

### Decomposition of NEET prevalence and penalty across periods

The decomposition of health exposures using 2014-2018 as the reference period as well as the FE estimates are shown in **Table 1**. The full results are in <u>supplementary files S4 and S5</u> <u>respectively</u>.

**Table 1.** Decomposition of the 16-24 NEET trends by health exposures comparing the 2009-13 and 2019-24 periods to 2014-18 and Fixed Effects interactions of exposure by time periods.

|  | 2009-13 to 2014-18 |  |  | 2019-23 to 2014-18 |  |  | FE<br>main effect<br>(2014-18) |
| --- | --- | --- | --- | --- | --- | --- | --- |
|  | Decomposition<br>Explained<br>component | Unexplained<br>component | FE<br>interaction | Decomposition<br>Explained<br>component | Unexplained<br>component | FE<br>interaction |  |
| Self-reported health conditions or disability | -0.215***<br>(-0.291;-0.140) | -2.718***<br>(-4.392; -1.043) | -0.148*<br>(-0.285;-0.011) | 0.581***<br>(0.425;0.740) | 0.350<br>(-2.077;2.777) | 0.068<br>(-0.096;0.232) | 0.105<br>(-0.002;0.213) |
| Psychological distress (GHQ-12 caseness) | -0.182***<br>(-0.248;-0.117) | -0.088<br>(-0.438;0.261) | -0.095<br>(-0.251;0.062) | 0.520***<br>(0.373;0.667) | 0.209<br>(-0.241;0.660) | 0.055<br>(-0.108;0.218) | 0.127*<br>(0.020;0.233) |
| Multimorbidity | 0.138***<br>(0.078;0.197) | -0.051<br>(-0.117;0.014) | 0.064<br>( -0.573;0.703) | 0.091*<br>(0.019;0.162) | 0.002<br>(-0.008;0.093) | -0.056<br>( -0.578; 0.465) | 0.205<br>( -0.118; 0.527) |
| Asthma | 0.074**<br>(0.015;0.134) | -0.055<br>(-0.361;0.251) | 0.232<br>(-0.031;0.495) | -0.018<br>(-0.047;0.012) | 0.117<br>(-0.497;0.264) | -0.227<br>(-0.529;0.075) | -0.092<br>(-0.538;0.354) |
| Depression | 0.125***<br>(0.070;0.181) | -0.065<br>(-0.159;0.029) | -0.012<br>(-0.314;0.288) | 0.472***<br>(0.307;0.637) | -0.019<br>(-0.129;0.089) | -0.177<br>(-0.478;0.123) | 0.326***<br>(0.119;0.532) |
| Diabetes | 0.011<br>(-0.005;0.028) | -0.007<br>(-0.044;0.031) | -0.371<br>(-0.980;0.238) | 0.018<br>(-0.008;0.044) | 0.003<br>(-0.041;0.047) | 0.106<br>(-0.840;1.052) | -0.105<br>(-0.868;0.658) |
| Epilepsy | 0.024*<br>(0.000;0.048) | 0.004<br>(-0.048;0.057) | -0.283<br>(-1.173;0.606) | 0.01<br>(-0.009;0.029) | -0.093*<br>(-0.167;-0.019) | -0.233<br>(-1.398;0.933) | 0.736<br>(-0.221;1.694) |
| High blood pressure | 0.053*<br>(0.008;0.098) | -0.049<br>(-0.110;0.011) | -0.024<br>(-0.958;0.911) | -0.063<br>(-0.120;-0.005) | (0.029<br>(-0.061;0.119) | -13.41***<br>(-14.87;-11.94) | -0.056<br>(-0.582;0.469) |

Note: \* p<0.05, \*\* p<0.01, \*\*\* p<0.001. Decomposition refers to Blinder-Oaxaca Probit decomposition comparing the periods 2009-13 and 2019-23 to the reference period (2014-18). FE refers to the Fixed Effects modified Poisson regression explaining NEET outcomes the interaction between the three periods (2014-18 as the reference) and the lagged exposure. The main effect represents the association with NEET at the reference period (2014-18) and the interaction terms refers to the change in coefficient associated with period effect. Self-reported health conditions or disability and psychological distress were collected at each wave. The other health conditions were collected at survey entry (except on wave 2) and refreshed based on newly diagnosed health conditions, they represent the diagnosis of health condition ever diagnosed except in the FE model that only accounts for change in diagnosed conditions.

Comparing 2009-2013 to 2014-2018, the explained component (prevalence) was negative for both self-reported health conditions or disability (-0.215, 95%CI -0.291,-0.140) and psychological distress (-0.182, 95%CI -0.248,-0.117). This indicates that the prevalence of these conditions increased over this period, which counteracted the decline in NEET rates. In other words, without these increases in ill health, the reduction in NEET between the two periods would have been larger. The unexplained component (penalty) was negative and significant for health conditions (-2.718, 95%CI -4.392,-1.043) but not significant for distress (-0.088, 95%CI -0.438; 0.261), indicating that the effect of general health conditions on NEET was weaker during the 2009-2013 period compared to 2014-18. The FE interaction terms confirm the causal nature of such a relationship with coefficients of respectively -0.148 (95%CI -0.285,-0.011) and -0.095 (95%CI -0.251,0.062).

Comparing 2019-2023 to 2014-2018, the explained component was positive and significant for health conditions or disability (0.581, 95%CI 0.425;0.740) and psychological distress (0.520, 95%CI 0.373,0.667). This means that higher prevalence of health problems in the 2019-2023 period pushed NEET rates up. The unexplained component and FE were not statistically significant.

For multimorbidity, comparing 2009-13 to 2014-18, the explained coefficient showed a positive contribution in the earlier period 0.138 (95%CI: 0.078;0.197). Comparing 2019-23 to the middle period, this contribution was also positive (0.091 95%CI: 0.019;0.162). But the composition of multimorbidity changed over time. On the one hand, asthma was a major contributor of NEET in the first period (0.074 95%: 0.015;0.134) but not in the latest. On the other hand, diagnosed depression explained higher NEET rates in the 2009-13 period (0.125 95%CI: 0.070;0.181) but the effect was higher in the 2019-23 period (0.472 95%CI 0.307;0.637). The unexplained and FE coefficients are not significant for these variables indicating no significant change in penalty. Epilepsy, diabetes, and high blood pressure were imprecise due to low prevalence.

**Figure 3** shows yearly BO estimates in comparison to year 2016 with full estimate in <u>supplementary file S7</u>. The figure illustrates the increasing contribution of self-reported health conditions or disability and psychological distress (explained component), the role of clinical depression in driving NEET increases in recent years, and the relatively stable penalty associated with these factors. The FE estimates – comparable to the unexplained component – show no difference as can be seen in <u>supplementary file S8</u>.

**Figure 3.**
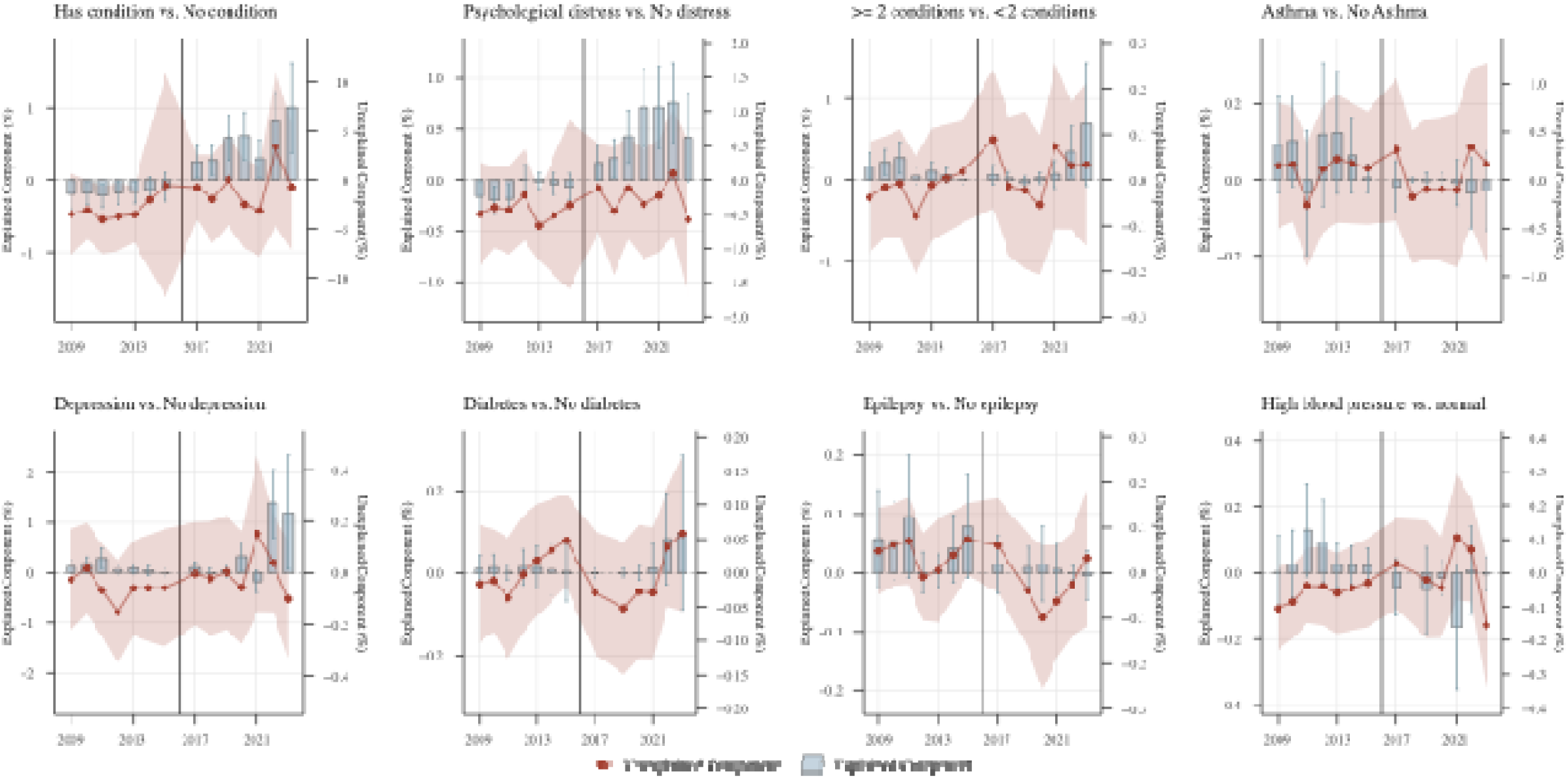
Decomposition of NEET trends comparing each year to the 2016-reference (probit)

Decomposition and FE results for socio-economic and demographic exposures are shown in <u>supplementary file S6</u>. Across high NEET periods, higher NEET rates were consistently linked to greater prevalence of structural disadvantages (low education, low household income, and deprivation) all of which showed positive and significant explained components. Child responsibilities also raised NEET in the earlier period, but in the latest period their prevalence had declined, pulling rates down instead. The unexplained component was largely stable across variables except age group and sex. The penalty for respondents aged 16-18, relative to the 19-24, increased sharply in the latest period (1.246 (95%CI: 0.795;1.696)), an effect confirmed by the fixed-effect interaction (0.367 (95%CI: 0.146,0.589)). By contrast, the penalty associated with being female reduced over the recent period -1.028 (95%CI: - 1.969;-0.086). These can be visualised in the model using the 2016 baseline reference in <u>supplementary file S7</u>.

Finally, as can be seen in supplementary files S4, the decomposition models include a constant unexplained term that is large across models, indicating that other factors contribute that explain NEET differences that are not captures by our models.

### Additional analyses

Three sets of supplementary analyses tested the robustness and sub-group differences of our main findings (<u>Supplementary files S9-S12</u>). First, adjusting simultaneously for both health conditions and psychological distress did not change the results. The explained component remained positive for both exposures when comparing 2019-23 to 2014-18 and negative when comparing 2009-13 to 2014-18. The unexplained components and FE went in a similar direction (negative in the earlier period and positive in the latest) but non-significant. Second, stratification by demographic and socio-economic variables shows no significant difference across sub-groups both in the BO decomposition (S10) and in the FE modelling (S11-12).

## Discussion

This study provides, to our knowledge, the first decomposition of rising NEET rates among young people into components attributable to changing health prevalence versus changing health penalty. Our findings highlight that the underlying issue of ill health – both physical and mental – within UK young people drives up NEET rates. Over the recent years, there was no higher NEET risks for those with poor health but rather an increased number of people with poor health explaining higher NEET rates.

Psychological distress and reporting a health condition or disability contributed to explaining NEET rates even when NEET was at its lowest (in 2014-18). Their increased prevalence despite falling NEET suggests these factors contributed to NEET, consistent with a causal role. It should be noted that increased ill health prevalence in the 16-24 population is not driven solely by the COVID-19 pandemic but started earlier and does not appear to correspond only to the effect of a one-time shock.^26^ The penalty associated with psychological distress remained stable over time. We observed a relative increase in penalty for those reporting a health condition or disability between the 2009-2013 and 2014-2018 but estimates have remained stable since then.

Clinical depression, by contrast, moved with NEET, rising when NEET rose and falling when NEET fell. This suggests it is more responsive to employment conditions and may be partly consequences of NEET rather than a driver. However, depression rates have more drastically increased in years 2022 and 2023 and contributed to higher NEET rates. The penalty associated with depression has not changed over time.

Multimorbidity contribution to NEET is more complex because conditions prevalences have changed over time. We observed declining asthma, consistent with other studies,^27^ but this was offset by rising diagnosed depressions, resulting in a net contribution that followed NEET.

These figures did not vary within socio-demographic subgroups but young age played a major role in explaining the recent NEET increase, with a substantially larger rise among 16- 17.

This study has several strengths. First, it uses data from a large-scale, nationally representative survey that provides robust sample sizes for analysing NEET trends among UK youth. Second, the cross-sectional decomposition approach, that is not commonly used in public health research, allows disentanglement of two distinct sources of change in NEET rates: prevalence and penalty. Third, the study benefits from rich information on a wide range of factors relevant to NEET risk, including health and socio-economic status.

Several limitations should be acknowledged. Although the fixed-effects modelling includes an exposure lag to address temporal ordering and strengthen causal inference, unmeasured and time varying confounding might influence our estimates. Early childhood exposures to poverty and socio-economic conditions are known to affect life course employment outcomes.^28,29^ While Understanding Society includes a youth module for respondents below age 15, including this information would drastically reduce the analytic sample and limit inference across periods. Similarly, information on obesity or BMI – which also contribute to non-employment ^30^ – is not sufficiently available. Blinder-Oaxaca decomposition does not accommodate modified Poisson regression. To approximate relative risks, we used probit modelling, which fits the distribution of the binary outcome and is the closest approximation to modified Poisson regression.

Finally, an unexplained component remained in our decomposition models, indicating that health and socio-demographic exposures do not fully capture the NEET trend. Other factors, including changes in educational engagement, economic uncertainty and contextual influences, may have contributed.

## Conclusion

To our knowledge, this study is the first to address whether rising NEET rates reflect increased ill health prevalence or increased risk of NEET among those with poor health. This distinction matters for policy. If rising NEET is driven by more young people having ill health, interventions should target mental health protection, early intervention, and treatment access. If rising NEET is driven by ill health becoming a stronger penalty, interventions should target workplace accommodations, anti-discrimination measures, employer practices and education access. Our findings suggest that the recent rise is driven predominantly by increased prevalence. This suggests that policymakers should continue to address labour market barriers for those with ill health but must also prioritise targeted interventions to improve both the physical and mental health of young people to tackle the NEET increase.

## Data Availability

Data are available from the UK Data Service (https://ukdataservice.ac.uk/) upon registration and application.

## Declarations

### Authors contributions

Conceptualization: JW, PP, NC; Methodology: JW, PP; Software: JW; Validation: ZL; Formal analysis: JW; Investigation: JW, PP; Resources: JW, PP, NC, GB; Data Curation: JW; Writing - Original Draft: JW; Writing - Review & Editing: DK, DS, CBS, ZL, GP, NC, PP; Visualization: JW; Supervision: JW, GB, PP, NC; Project administration: JW, PP; Funding acquisition: PP, NC, GP, JW.

