## Supplementary files S1-S12 for "Rising rates of young people Not in Education, Employment, or Training (NEET) explained by higher prevalence of physical and psychological ill health: a 15-year UK study"

### Supplementary file S1. Sample composition

| Core variables |  | Full sample | |  | Lag exposure sample | |
| --- | --- | --- | --- | --- | --- | --- |
|  |  | Observations | Respondents |  | Observations | Respondents |
| **NEET** | | 66,160 | 15,242 |  |  |  |
| Health conditions or disability | | 65,968 | 15,242 |  | 46,153 | 14,616 |
|  | *Proportion of original sample* |  |  |  | *0.700* | *0.959* |
| **GHQ-12 caseness** | | 56,242 | 14,429 |  | 39,963 | 13,314 |
|  | *Proportion of original sample* |  |  |  | *0.711* | *0.923* |
| **Health conditions & multimorbidity** | | 49,932 | 12,147 |  | 35,131 | 11,415 |
|  | *Proportion of original sample* |  |  |  | *0.704* | *0.940* |

### Supplementary file S2. Probability of no two consecutive waves in a row by NEET Status, year, self-reported health condition or disability, GHQ-12 caseness, age group, sex, ethnicity, education, region, child responsibility, equivalized household incomes and IMD

|  | Probit | Robust Standard Errors | P-value | [95% conf. interval] | |
| --- | --- | --- | --- | --- | --- |
| NEET | 0.019 | 0.017 | 0.266 | -0.015 | 0.053 |
| year |  |  |  |  |  |
| 2010 | -0.189 | 0.035 | 0.000 | -0.257 | -0.121 |
| 2011 | -0.118 | 0.034 | 0.001 | -0.185 | -0.051 |
| 2012 | 0.035 | 0.034 | 0.307 | -0.032 | 0.102 |
| 2013 | 0.136 | 0.034 | 0.000 | 0.069 | 0.203 |
| 2014 | 0.280 | 0.034 | 0.000 | 0.212 | 0.347 |
| 2015 | 0.270 | 0.034 | 0.000 | 0.202 | 0.337 |
| 2016 | 0.303 | 0.034 | 0.000 | 0.235 | 0.370 |
| 2017 | 0.286 | 0.035 | 0.000 | 0.217 | 0.355 |
| 2018 | 0.279 | 0.036 | 0.000 | 0.209 | 0.348 |
| 2019 | 0.230 | 0.036 | 0.000 | 0.160 | 0.300 |
| 2020 | 0.162 | 0.037 | 0.000 | 0.091 | 0.234 |
| 2021 | -0.016 | 0.038 | 0.673 | -0.091 | 0.058 |
| 2022 | -0.082 | 0.040 | 0.042 | -0.160 | -0.003 |
| 2023 | 0.143 | 0.051 | 0.005 | 0.043 | 0.243 |
| Self-reported health conditions or disability | -0.021 | 0.016 | 0.175 | -0.051 | 0.009 |
| Psychological distress (GHQ-12 caseness) | 0.004 | 0.013 | 0.728 | -0.020 | 0.029 |
| Age group (17-18, ref.: 19-24) | 0.024 | 0.014 | 0.101 | -0.005 | 0.052 |
| Sex (female, ref.: male) | -0.004 | 0.011 | 0.702 | -0.026 | 0.018 |
| Ethnicity (non-white, ref.: white) | -0.080 | 0.013 | 0.000 | -0.106 | -0.055 |
| Education (max. GCSE) | 0.029 | 0.013 | 0.024 | 0.004 | 0.054 |
| Region (London & South of England, ref.: other) | -0.073 | 0.011 | 0.000 | -0.094 | -0.052 |
| Child responsibilities (yes, ref.: no child responsibilities) | -0.212 | 0.027 | 0.000 | -0.264 | -0.160 |
| Lowest household incomes quintiles | 0.011 | 0.014 | 0.428 | -0.016 | 0.039 |
| Multiple deprivation lowest quintile | 0.024 | 0.013 | 0.063 | -0.001 | 0.050 |
| Constant | -0.116 | 0.047 | 0.013 | -0.207 | -0.025 |

### Supplementary file S3. Prevalence and risk ratios of socio-economic variables

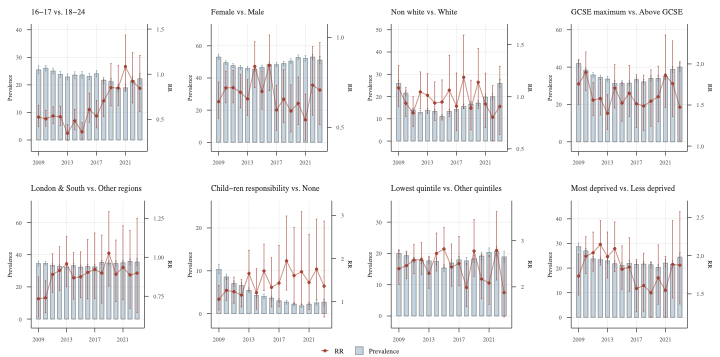

### Supplementary file S4. Blinder-Oaxaca decomposition full estimates (2009-13 to 2014-18 and 2019-2023 to 2014-18), probit

**2009-13 to 2014-18**

|  | **N** | **Total Gap** | **95%CI** | | **Explained** | **95%CI** | | **Unexplained** | **95%CI** | |
| --- | --- | --- | --- | --- | --- | --- | --- | --- | --- | --- |
| Self-reported health conditions or disability | 50.072 | 0.02083 | 0.01392 | 0.02773 | -0.00216 | -0.00291 | -0.00140 | 0.02298 | 0.01607 | 0.02989 |
| Psychological distress (GHQ-12 caseness) | 50.164 | 0.01893 | 0.01177 | 0.02608 | -0.00182 | -0.00248 | -0.00117 | 0.02076 | 0.01358 | 0.02793 |
| Multimorbidity | 38.435 | 0.02776 | 0.02000 | 0.03553 | 0.00138 | 0.00078 | 0.00197 | -0.00518 | -0.00117 | 0.00014 |
| Asthma | 38.435 | 0.02776 | 0.02000 | 0.03553 | 0.00075 | 0.00016 | 0.00134 | 0.02702 | 0.01926 | 0.03477 |
| Depression | 38.434 | 0.02776 | 0.02000 | 0.03553 | 0.00125 | 0.00070 | 0.00181 | -0.00065 | -0.00159 | 0.00029 |
| Diabetes | 38.435 | 0.02776 | 0.02000 | 0.03553 | 0.00011 | -0.00005 | 0.00028 | 0.02765 | 0.01989 | 0.03541 |
| Epilepsy | 38.435 | 0.02776 | 0.02000 | 0.03553 | 0.00024 | 0.00000 | 0.00048 | 0.02752 | 0.01977 | 0.03528 |
| High blood pressure | 48.435 | 0.02776 | 0.02000 | 0.03552 | 0.00053 | 0.00008 | 0.00098 | 0.02723 | 0.01948 | 0.03499 |
| Age group (17-18, ref.: 19-24) | 50.164 | 0.02071 | 0.01377 | 0.02764 | -0.00111 | -0.00187 | -0.00035 | 0.02182 | 0.01489 | 0.02875 |
| Sex (female, ref.: male) | 50.164 | 0.02071 | 0.01378 | 0.02764 | -0.00032 | -0.00082 | 0.00017 | 0.02103 | 0.01410 | 0.02796 |
| Ethnicity (non-white, ref.: white) | 48.193 | 0.02220 | 0.01516 | 0.02925 | -0.00007 | -0.00044 | 0.00031 | 0.02227 | 0.01517 | 0.02937 |
| Education (max. GCSE) | 49.288 | 0.02434 | 0.01741 | 0.03127 | 0.00316 | 0.00237 | 0.00395 | 0.02118 | 0.01430 | 0.02805 |
| Region (London & South of England, ref.: other) | 50.136 | 0.02083 | 0.01390 | 0.02776 | -0.00008 | -0.00031 | 0.00014 | 0.02092 | 0.01399 | 0.02784 |
| Child responsibilities (yes, ref.: no child responsibilities) | 49.383 | 0.01994 | 0.01297 | 0.02691 | 0.00128 | 0.00059 | 0.00196 | 0.01866 | 0.01166 | 0.02567 |
| Lowest household incomes quintiles | 49.505 | 0.02041 | 0.01346 | 0.02736 | 0.00206 | 0.00107 | 0.00305 | 0.01835 | 0.01148 | 0.02522 |
| Multiple deprivation lowest quintile | 50.136 | 0.02083 | 0.01392 | 0.02775 | 0.00350 | 0.00261 | 0.00439 | 0.01733 | 0.01049 | 0.02417 |
|  | **N** | **Variable explained** | **95%CI** | | **Variable unexplained** | **95%CI** | | **Constant** | **95%CI** | |
| Self-reported health conditions or disability | 50.072 | -0.00216 | -0.00291 | -0.00140 | -0.02718 | -0.04392 | -0.01043 | 0.05016 | 0.03235 | 0.06797 |
| Psychological distress (GHQ-12 caseness) | 50.164 | -0.00182 | -0.00248 | -0.00117 | -0.00088 | -0.00438 | 0.00261 | 0.02164 | 0.01381 | 0.02947 |
| Multimorbidity | 38.435 | 0.00086 | 0.00034 | 0.00139 | 0.00009 | -0.00031 | 0.00050 | 0.02679 | 0.01901 | 0.03458 |
| Asthma | 38.435 | 0.00075 | 0.00016 | 0.00134 | -0.00055 | -0.00361 | 0.00251 | 0.02757 | 0.01914 | 0.03600 |
| Hypothyroidism | 38.434 | -0.00004 | -0.00013 | 0.00006 | 0.00027 | -0.00013 | 0.00068 | 0.02753 | 0.01975 | 0.03531 |
| Diabetes | 38.435 | 0.00011 | -0.00005 | 0.00028 | -0.00007 | -0.00044 | 0.00031 | 0.02772 | 0.01994 | 0.03549 |
| Epilepsy | 38.435 | 0.00024 | 0.00000 | 0.00048 | 0.00004 | -0.00048 | 0.00057 | 0.02748 | 0.01969 | 0.03527 |
| High blood pressure | 48.435 | 0.00053 | 0.00008 | 0.00098 | -0.00049 | -0.00110 | 0.00011 | 0.02773 | 0.01995 | 0.03551 |
| Age group (17-18, ref.: 19-24) | 50.164 | -0.00111 | -0.00187 | -0.00035 | -0.00292 | -0.00747 | 0.00163 | 0.02473 | 0.01621 | 0.03326 |
| Sex (female, ref.: male) | 50.164 | -0.00032 | -0.00082 | 0.00017 | -0.00314 | -0.00999 | 0.00370 | 0.02417 | 0.01371 | 0.03463 |
| Ethnicity (non-white, ref.: white) | 48.193 | -0.00007 | -0.00044 | 0.00031 | -0.00051 | -0.00302 | 0.00201 | 0.02278 | 0.01498 | 0.03058 |
| Education (max. GCSE) | 49.288 | 0.00316 | 0.00237 | 0.00395 | 0.00265 | -0.00217 | 0.00747 | 0.01853 | 0.01009 | 0.02696 |
| Region (London & South of England, ref.: other) | 50.136 | -0.00008 | -0.00031 | 0.00014 | -0.00246 | -0.00767 | 0.00275 | 0.02337 | 0.01492 | 0.03183 |
| Child responsibilities (yes, ref.: no child responsibilities) | 49.383 | 0.00128 | 0.00059 | 0.00196 | -0.00067 | -0.00167 | 0.00033 | 0.01934 | 0.01218 | 0.02649 |
| Lowest household incomes quintiles | 49.505 | 0.00206 | 0.00107 | 0.00305 | 0.00104 | -0.00182 | 0.00390 | 0.01731 | 0.00987 | 0.02475 |
| Multiple deprivation lowest quintile | 50.136 | 0.00350 | 0.00261 | 0.00439 | 0.00429 | 0.00040 | 0.00819 | 0.01304 | 0.00496 | 0.02111 |

**2019-23 to 2013-18**

|  | **N** | **Total Gap** | **95%CI** | | **Explained** | **95%CI** | | **Unexplained** | **95%CI** | |
| --- | --- | --- | --- | --- | --- | --- | --- | --- | --- | --- |
| Self-reported health conditions or disability | 37.684 | 0.03063 | 0.02136 | 0.03991 | 0.00582 | 0.00425 | 0.00739 | 0.02481 | 0.01578 | 0.03385 |
| Psychological distress (GHQ-12 caseness) | 37.867 | 0.03069 | 0.02122 | 0.04017 | 0.00520 | 0.00373 | 0.00487 | 0.02539 | 0.01617 | 0.03481 |
| Multimorbidity | 34.357 | 0.03599 | 0.02645 | 0.04554 | 0.00091 | 0.00019 | 0.00162 | 0.00002 | -0.00008 | 0.00093 |
| Asthma | 34.357 | 0.03599 | 0.02645 | 0.04554 | -0.00018 | -0.00047 | 0.00012 | 0.03617 | 0.02662 | 0.04572 |
| Depression | 34.357 | 0.03599 | 0.02646 | 0.04553 | 0.00472 | 0.00307 | 0.00637 | -0.00019 | -0.00129 | 0.00089 |
| Diabetes | 34.357 | 0.03599 | 0.02645 | 0.04553 | 0.00018 | -0.00008 | 0.00044 | 0.03581 | 0.02628 | 0.04535 |
| Epilepsy | 34.357 | 0.03599 | 0.02645 | 0.04554 | 0.00010 | -0.00009 | 0.00029 | 0.03589 | 0.02636 | 0.04543 |
| High blood pressure | 34.357 | 0.03599 | 0.02646 | 0.04553 | -0.00063 | -0.00120 | -0.00005 | 0.03662 | 0.02706 | 0.04618 |
| Age group (17-18, ref.: 19-24) | 37.867 | 0.03079 | 0.02147 | 0.04011 | 0.00039 | -0.00012 | 0.00091 | 0.03040 | 0.02112 | 0.03968 |
| Sex (female, ref.: male) | 37.866 | 0.03079 | 0.02149 | 0.04010 | -0.00283 | -0.00392 | -0.00174 | 0.03362 | 0.02416 | 0.04308 |
| Ethnicity (non-white, ref.: white) | 37.357 | 0.03184 | 0.02250 | 0.04119 | -0.00037 | -0.00137 | 0.00062 | 0.03222 | 0.02273 | 0.04171 |
| Education (max. GCSE) | 37.374 | 0.03245 | 0.02314 | 0.04176 | 0.00285 | 0.00175 | 0.00395 | 0.02960 | 0.02041 | 0.03879 |
| Region (London & South of England, ref.: other) | 37.848 | 0.03081 | 0.02149 | 0.04012 | -0.00020 | -0.00053 | 0.00014 | 0.03100 | 0.02168 | 0.04033 |
| Child responsibilities (yes, ref.: no child responsibilities) | 37.867 | 0.03079 | 0.02147 | 0.04011 | -0.00104 | -0.00182 | -0.00025 | 0.03183 | 0.02246 | 0.04119 |
| Lowest household incomes quintiles | 36.189 | 0.03005 | 0.02054 | 0.03957 | 0.00353 | 0.00208 | 0.00497 | 0.02653 | 0.01721 | 0.03585 |
| Multiple deprivation lowest quintile | 37.846 | 0.03082 | 0.02151 | 0.04013 | 0.00004 | -0.00079 | 0.00088 | 0.03078 | 0.02151 | 0.04005 |
|  | **N** | **Variable explained** | **95%CI** | | **Variable unexplained** | **95%CI** | | **Constant** | **95%CI** | |
| Self-reported health conditions or disability | 37.684 | 0.00582 | 0.00425 | 0.00739 | 0.00350 | -0.02077 | 0.02777 | 0.02131 | -0.00463 | 0.04726 |
| Psychological distress (GHQ-12 caseness) | 37.867 | 0.00520 | 0.00373 | 0.00667 | 0.00209 | -0.00241 | 0.00660 | 0.02339 | 0.01279 | 0.03400 |
| Multimorbidity | 34.357 | -0.00039 | -0.00078 | 0.00000 | 0.00045 | -0.00016 | 0.00107 | 0.03592 | 0.02636 | 0.04549 |
| Asthma | 34.357 | -0.00018 | -0.00047 | 0.00012 | -0.00117 | -0.00497 | 0.00264 | 0.03734 | 0.02708 | 0.04759 |
| Hypothyroidism | 34.357 | 0.00020 | -0.00014 | 0.00053 | 0.00037 | -0.00011 | 0.00086 | 0.03542 | 0.02589 | 0.04495 |
| Diabetes | 34.357 | 0.00018 | -0.00008 | 0.00044 | 0.00003 | -0.00041 | 0.00047 | 0.03578 | 0.02623 | 0.04534 |
| Epilepsy | 34.357 | 0.00010 | -0.00009 | 0.00029 | -0.00093 | -0.00167 | -0.00019 | 0.03682 | 0.02723 | 0.04641 |
| High blood pressure | 34.357 | -0.00063 | -0.00120 | -0.00005 | 0.00029 | -0.00061 | 0.00119 | 0.03633 | 0.02679 | 0.04588 |
| Age group (17-18, ref.: 19-24) | 37.867 | 0.00039 | -0.00012 | 0.00091 | 0.01246 | 0.00795 | 0.01696 | 0.01794 | 0.00781 | 0.02807 |
| Sex (female, ref.: male) | 37.866 | -0.00283 | -0.00392 | -0.00174 | -0.01028 | -0.01969 | -0.00086 | 0.04390 | 0.02941 | 0.05839 |
| Ethnicity (non-white, ref.: white) | 37.357 | -0.00037 | -0.00137 | 0.00062 | -0.00111 | -0.00426 | 0.00203 | 0.03333 | 0.02288 | 0.04378 |
| Education (max. GCSE) | 37.374 | 0.00285 | 0.00175 | 0.00395 | 0.00329 | -0.00289 | 0.00947 | 0.02631 | 0.01523 | 0.03740 |
| Region (London & South of England, ref.: other) | 37.848 | -0.00020 | -0.00053 | 0.00014 | 0.00132 | -0.00514 | 0.00779 | 0.02968 | 0.01830 | 0.04106 |
| Child responsibilities (yes, ref.: no child responsibilities) | 37.867 | -0.00104 | -0.00182 | -0.00025 | 0.00048 | -0.00121 | 0.00218 | 0.03134 | 0.02190 | 0.04078 |
| Lowest household incomes quintiles | 36.189 | 0.00353 | 0.00208 | 0.00497 | 0.00010 | -0.00346 | 0.00366 | 0.02643 | 0.01648 | 0.03638 |
| Multiple deprivation lowest quintile | 37.846 | 0.00004 | -0.00079 | 0.00088 | -0.00041 | -0.00479 | 0.00398 | 0.03119 | 0.02107 | 0.04130 |

### Supplementary file S5. Fixed effects Poisson regression estimates

|  | **Health** | **Psychological distress** | **Multimorbidity** | **Asthma** | **Depression** | **Diabetes** |
| --- | --- | --- | --- | --- | --- | --- |
| Period 2009-2013 | -0.021 | -0.005 | -0.036 | -0.084 | -0.037 | -0.033 |
|  | (-0.107,0.065) | (-0.105,0.095) | (-0.142, 0.068) | (-0.201,0.033) | (-0.136,0.060) | (-0.140,0.073) |
| Period 2019-2023 | 0.329^***^ | 0.300^***^ | 0.334^***^ | 0.365^***^ | 0.367^***^ | 0.333^***^ |
|  | (0.213,0.444) | (0.180,0.421) | (0.229,0.439) | (0.254,0.476) | (0.263,0.470) | (0.229,0.437) |
| Variable | 0.105 | 0.127^*^ | 0.205 | -0.092 | 0.326 | -0.105 |
|  | (-0.002,0.213) | (0.020,0.233) | (-0.118,0.527) | (-0.538,0.354) | (0.119,0.532) | (-0.868,0.658) |
| Variable * Period 2009-2013 | -0.148^*^ | -0.095 | 0.064 | 0.232 | -0.012 | -0.371 |
|  | (-0.285,-0.011) | (-0.251,0.062) | (-0.573,0.702) | (-0.031,0.495) | (-0.314, 0.288) | (-0.980,0.238) |
| Variable * Period 2019-2023 | 0.068 | 0.055 | -0.056 | -0.227 | -0.177 | 0.106 |
|  | (-0.096,0.232) | (-0.108,0.218) | (-0.578,0.465) | (-0.529,0.075) | (-0.478,0.123) | (-0.840,1.052) |
| N | 13326 | 10745 | 9837 | 9837 | 9837 | 9837 |
|  | **Epilepsy** | **High blood pressure** | **Age group** | **Child responsibility** | **Incomes** | **IMD** |
| Period 2009-2013 | -0.034 | -0.038 | 0.099^*^ | -0.065 | -0.053 | -0.081 |
|  | (-0.140,0.071) | (-0.143,0.067) | (0.015,0.184) | (-0.151,0.020) | (-0.141,0.035) | (-0.178,0.015) |
| Period 2019-2023 | 0.334^***^ | 0.337^***^ | 0.223^***^ | 0.348^***^ | 0.304^***^ | 0.305^***^ |
|  | (0.230,0.438) | (0.233,0.442) | (0.114,0.332) | (0.244,0.451) | (0.190,0.419) | (0.177,0.433) |
| Variable | 0.736 | -0.056 | -0.253^***^ | -0.162 | 0.013 | 0.004 |
|  | (-0.221,1.694) | (-0.582,0.469) | (-0.380,-0.126) | (-0.409,0.085) | (-0.078,0.104) | (-0.189,0.198) |
| Variable * Period 2009-2013 | -0.283 | -0.024 | -0.146 | 0.208 | -0.007 | 0.054 |
|  | (-1.173,0.606) | (-0.958,0.911) | (-0.313,0.022) | (-0.099,0.515) | (-0.135,0.120) | (-0.097,0.204) |
| Variable * Period 2019-2023 | -0.233 | -13.409^***^ | 0.367^**^ | -0.154 | 0.124 | 0.14 |
|  | (-1.398,0.933) | (-14.874,-11.945) | (0.146,0.589) | (-0.555,0.247) | (-0.029,0.277) | (-0.049,0.329) |
| N | 9837 | 9837 | 13381 | 11923 | 13007 | 13368 |

### Supplementary file S6. Decomposition of the 16-24 NEET trends by demographic and socioeconomic factors comparing the 2009-13 and 2019-24 periods to 2014-18 and Fixed Effects interactions of exposure by time periods.

|  | **2009-13 to 2014-18** | | |  | **2019-23 to 2014-18** | | |  | FE  main effect  (2014-18) |
| --- | --- | --- | --- | --- | --- | --- | --- | --- | --- |
|  | Decomposition | | FE  interaction |  | Decomposition | | FE  interaction |  |  |
|  | Explained component | Unexplained component |  |  | Explained component | Unexplained component |  |  |  |
| Age group (16-18, ref.: 19-24) | -0.111^***^ | -0.292 | -0.146 |  | 0.039 | 1.246^***^ | 0.367^**^ |  | -0.253^***^ |
|  | (-0.187;-0.035) | (-0.747;0.163) | (-0.313,0.022) |  | (-0.012;0.091) | (0.795;1.696) | (0.146,0.589) |  | (-0.380,-0.126) |
| Sex (female, ref.: male) | -0.032 | -0.314 |  |  | -0.283^***^ | -1.028^*^ |  |  |  |
|  | (-0.082;0.017) | (-0.999;0.370) |  |  | (-0.392;-0.174) | (-1.969;-0.086) |  |  |  |
| Ethnicity (non-white, ref.: white) | -0.007 | -0.051 |  |  | -0.037 | -0.111 |  |  |  |
|  | (-0.044;0.031) | (-0.302;0.201) |  |  | (-0.137;0.062) | (-0.426;0.203) |  |  |  |
| Education (max. GCSE) | 0.316^***^ | 0.265 |  |  | 0.285^***^ | 0.329 |  |  |  |
|  | (0.237;0.395) | (-0.217;0.747) |  |  | (0.175;0.395) | (-0.289;0.947) |  |  |  |
| Region (London & South of England, ref.: other) | -0.008 | -0.246 |  |  | -0.02 | 0.132 |  |  |  |
|  | (-0.031;0.014) | (-0.767;0.275) |  |  | (-0.053;0.014) | (-0.514;0.779) |  |  |  |
| Child responsibilities (yes, ref.: none) | 0.128^***^ | -0.067 | 0.208 |  | -0.104^**^ | 0.048 | -0.154 |  | -0.162 |
|  | (0.059;0.196) | (-0.167;0.033) | (-0.099,0.515) |  | (-0.182;-0.025) | (-0.121;0.218) | (-0.555,0.247) |  | (-0.409,0.085) |
| Lowest household incomes quintiles | 0.206^***^ | 0.104 | -0.007 |  | 0.353^***^ | 0.01 | 0.124 |  | 0.013 |
|  | (0.107;0.305) | (-0.182;0.390) | (-0.135,0.120) |  | (0.208;0.497) | (-0.346;0.366) | (-0.029,0.277) |  | (-0.078,0.104) |
| Multiple deprivation lowest quintile | 0.350^***^ | 0.429^*^ | 0.054 |  | 0.004 | -0.041 | 0.14 |  | 0.004 |
|  | (0.261;0.439) | (0.040;0.819) | (-0.097,0.204) |  | (-0.079;0.088) | (-0.479;0.398) | (-0.049,0.329) |  | (-0.189,0.198) |

Note: * p<0.05, ** p<0.01, *** p<0.001. Decomposition refers to Blinder-Oaxaca Probit decomposition comparing the periods 2009-13 and 2019-23 to the reference period (2014-18). FE refers to the Fixed Effects modified Poisson regression explaining NEET outcomes the interaction between the three periods (2014-18 as the reference) and the lagged exposure.. The main effect represents the association with NEET at the reference period (2014-18) and the interaction terms refers to the change in coefficient associated with period effect. All variables were collected at each wave

### Supplementary file S7. Blinder Oaxaca Decomposition using 2016 as the baseline reference (explained and unexplained components and 95%CI) – probit

**Health variables**

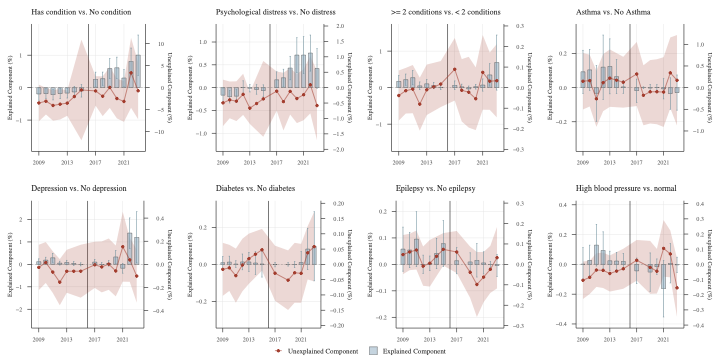

**Socio-economic variables**

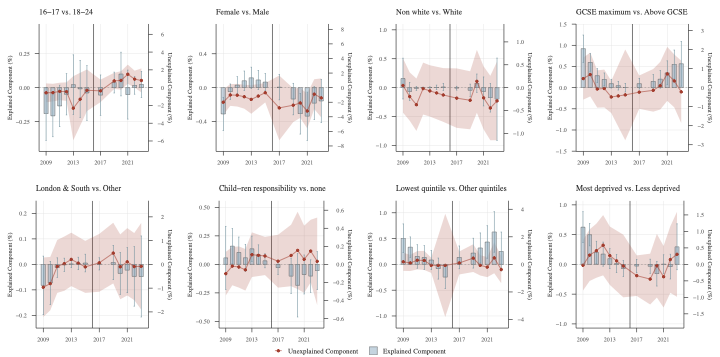

|  |  | Explained | | | Unexplained | | |
| --- | --- | --- | --- | --- | --- | --- | --- |
| Variable | Year | PP | 95%CI lower | 95%CI upper | PP | 95%CI lower | 95%CI upper |
| >= 2 conditions vs. < 2 conditions | 2009 | 0.0017248 | 0.0001243 | 0.0033253 | -0.0003839 | -0.0016027 | 0.0008349 |
| >= 2 conditions vs. < 2 conditions | 2010 | 0.0021944 | 0.0006598 | 0.0037291 | -0.0001496 | -0.0012594 | 0.0009602 |
| >= 2 conditions vs. < 2 conditions | 2011 | 0.0027342 | 0.000768 | 0.0047004 | -0.0000714 | -0.0012448 | 0.001102 |
| >= 2 conditions vs. < 2 conditions | 2012 | 0.0004917 | -0.0005803 | 0.0015636 | -0.0008037 | -0.0020929 | 0.0004854 |
| >= 2 conditions vs. < 2 conditions | 2013 | 0.0010521 | -0.0001035 | 0.0022076 | -0.0001041 | -0.0013317 | 0.0011235 |
| >= 2 conditions vs. < 2 conditions | 2014 | 0.0007531 | -0.0002889 | 0.0017952 | -0.0002103 | -0.0021754 | 0.0017548 |
| >= 2 conditions vs. < 2 conditions | 2015 | 0.0000758 | -0.0005199 | 0.0006716 | 0.0001738 | -0.0009448 | 0.0012924 |
| >= 2 conditions vs. < 2 conditions | 2017 | 0.0006594 | -0.0005176 | 0.0018363 | 0.0008934 | -0.0006273 | 0.0024141 |
| >= 2 conditions vs. < 2 conditions | 2018 | 0.0002336 | -0.0004962 | 0.0009634 | -0.0001294 | -0.0016465 | 0.0013877 |
| >= 2 conditions vs. < 2 conditions | 2019 | -0.0003459 | -0.0011407 | 0.0004489 | -0.0002226 | -0.0019447 | 0.0014994 |
| >= 2 conditions vs. < 2 conditions | 2020 | 0.000257 | -0.0003898 | 0.0009039 | -0.00054 | -0.0020654 | 0.0009854 |
| >= 2 conditions vs. < 2 conditions | 2021 | 0.0007126 | -0.0009509 | 0.0023761 | 0.0007427 | -0.0010743 | 0.0025597 |
| >= 2 conditions vs. < 2 conditions | 2022 | 0.0034866 | 0.0004208 | 0.0065523 | 0.0003133 | -0.0010789 | 0.0017055 |
| >= 2 conditions vs. < 2 conditions | 2023 | 0.0068711 | -0.0006688 | 0.014411 | 0.0003363 | -0.0014446 | 0.0021172 |
| 16-17 vs. 18-24 | 2009 | -0.0019206 | -0.0038701 | 0.0000289 | -0.0058029 | -0.0163848 | 0.004779 |
| 16-17 vs. 18-24 | 2010 | -0.0020726 | -0.0036157 | -0.0005295 | -0.0056141 | -0.0155601 | 0.0043319 |
| 16-17 vs. 18-24 | 2011 | -0.0013447 | -0.002873 | 0.0001837 | -0.0042073 | -0.0139702 | 0.0055556 |
| 16-17 vs. 18-24 | 2012 | -0.0004918 | -0.0020081 | 0.0010244 | -0.0044961 | -0.0149278 | 0.0059357 |
| 16-17 vs. 18-24 | 2013 | 0.0002208 | -0.0019679 | 0.0024096 | -0.0231417 | -0.0591863 | 0.0129028 |
| 16-17 vs. 18-24 | 2014 | -0.0002745 | -0.0017167 | 0.0011677 | -0.002367 | -0.0167728 | 0.0120388 |
| 16-17 vs. 18-24 | 2015 | -0.0003933 | -0.0024048 | 0.0016181 | -0.0029781 | -0.0264059 | 0.0204497 |
| 16-17 vs. 18-24 | 2017 | -0.0005495 | -0.0020314 | 0.0009324 | -0.0036198 | -0.0162657 | 0.0090261 |
| 16-17 vs. 18-24 | 2019 | 0.0003749 | -0.000394 | 0.0011438 | 0.0075855 | -0.0015019 | 0.0166729 |
| 16-17 vs. 18-24 | 2020 | 0.0010004 | -0.0006089 | 0.0026097 | 0.0080738 | -0.0023114 | 0.018459 |
| 16-17 vs. 18-24 | 2021 | -0.0005094 | -0.0023121 | 0.0012933 | 0.0150695 | 0.0042589 | 0.0258802 |
| 16-17 vs. 18-24 | 2022 | 0.0001671 | -0.0004814 | 0.0008157 | 0.0095511 | 8.29E-06 | 0.0190939 |
| 16-17 vs. 18-24 | 2023 | 0.0002349 | -0.0007305 | 0.0012003 | 0.0080787 | -0.0054489 | 0.0216064 |
| Asthma vs. No Asthma | 2009 | 0.0009304 | -0.0003319 | 0.0021927 | 0.0014792 | -0.005037 | 0.0079954 |
| Asthma vs. No Asthma | 2010 | 0.0010422 | -0.0001552 | 0.0022396 | 0.0016232 | -0.0043557 | 0.0076022 |
| Asthma vs. No Asthma | 2011 | -0.0003353 | -0.0019668 | 0.0012961 | -0.0025719 | -0.0090514 | 0.0039076 |
| Asthma vs. No Asthma | 2012 | 0.0011983 | -0.000687 | 0.0030837 | 0.0011547 | -0.0054324 | 0.0077418 |
| Asthma vs. No Asthma | 2013 | 0.0012466 | -0.000365 | 0.0028581 | 0.0021898 | -0.0044338 | 0.0088135 |
| Asthma vs. No Asthma | 2014 | 0.0011198 | -0.0001404 | 0.0023799 | 0.0022683 | -0.0073504 | 0.011887 |
| Asthma vs. No Asthma | 2015 | 0.0000456 | -0.0003288 | 0.0004199 | 0.0012503 | -0.0046188 | 0.0071193 |
| Asthma vs. No Asthma | 2017 | -0.000179 | -0.0008594 | 0.0005013 | 0.0031469 | -0.0042561 | 0.0105498 |
| Asthma vs. No Asthma | 2018 | -1.33E-06 | -0.0000479 | 0.0000453 | -0.0017671 | -0.0086924 | 0.0051583 |
| Asthma vs. No Asthma | 2019 | -0.000023 | -0.0002324 | 0.0001863 | -0.0009582 | -0.0082735 | 0.0063571 |
| Asthma vs. No Asthma | 2020 | 0.0000125 | -0.0001463 | 0.0001713 | -0.0009464 | -0.0084496 | 0.0065568 |
| Asthma vs. No Asthma | 2021 | -0.0000684 | -0.0006615 | 0.0005248 | -0.0010251 | -0.0090744 | 0.0070242 |
| Asthma vs. No Asthma | 2022 | -0.0003449 | -0.0012636 | 0.0005739 | 0.0034142 | -0.0047522 | 0.0115805 |
| Asthma vs. No Asthma | 2023 | -0.0002884 | -0.0013188 | 0.0007419 | 0.0016981 | -0.0087476 | 0.0121439 |
| Child-ren responsibility vs. none | 2009 | 0.0005727 | -0.0021979 | 0.0033433 | -0.0010289 | -0.0034458 | 0.001388 |
| Child-ren responsibility vs. none | 2010 | 0.0015961 | 0.0000469 | 0.0031453 | -0.0001937 | -0.0023134 | 0.001926 |
| Child-ren responsibility vs. none | 2011 | 0.001044 | -0.0003129 | 0.002401 | -0.0002906 | -0.0025716 | 0.0019903 |
| Child-ren responsibility vs. none | 2012 | 0.0005813 | -0.0006017 | 0.0017644 | -0.0006006 | -0.0029007 | 0.0016994 |
| Child-ren responsibility vs. none | 2013 | 0.0013675 | 0.0002492 | 0.0024858 | 0.0010729 | -0.0014002 | 0.003546 |
| Child-ren responsibility vs. none | 2014 | 0.000143 | -0.0002475 | 0.0005335 | -0.0004231 | -0.0031353 | 0.0022892 |
| Child-ren responsibility vs. none | 2015 | 0.0003171 | -0.0003038 | 0.0009381 | 0.00093 | -0.0011858 | 0.0030457 |
| Child-ren responsibility vs. none | 2017 | -0.0002938 | -0.000894 | 0.0003065 | 0.0003529 | -0.0025744 | 0.0032801 |
| Child-ren responsibility vs. none | 2019 | -0.001033 | -0.0025649 | 0.0004989 | 0.000987 | -0.0024192 | 0.0043932 |
| Child-ren responsibility vs. none | 2020 | -0.001838 | -0.0046143 | 0.0009383 | 0.0015479 | -0.0027325 | 0.0058283 |
| Child-ren responsibility vs. none | 2021 | -0.000886 | -0.0025977 | 0.0008256 | 0.0005638 | -0.0031574 | 0.004285 |
| Child-ren responsibility vs. none | 2022 | -0.0010272 | -0.0024508 | 0.0003965 | 0.0014747 | -0.0019941 | 0.0049434 |
| Child-ren responsibility vs. none | 2023 | -0.0005506 | -0.0022137 | 0.0011124 | 0.0003352 | -0.0045191 | 0.0051895 |
| Depression vs. No depression | 2009 | 0.0011973 | -0.0000605 | 0.0024551 | -0.0002635 | -0.0022249 | 0.0016979 |
| Depression vs. No depression | 2010 | 0.0018933 | 0.0005767 | 0.00321 | 0.0001629 | -0.0016047 | 0.0019306 |
| Depression vs. No depression | 2011 | 0.002938 | 0.000949 | 0.004927 | -0.0006642 | -0.0024628 | 0.0011343 |
| Depression vs. No depression | 2012 | 0.0005706 | -0.0002967 | 0.0014379 | -0.0015391 | -0.003535 | 0.0004568 |
| Depression vs. No depression | 2013 | 0.0008204 | -0.0002043 | 0.0018451 | -0.0005954 | -0.0024534 | 0.0012625 |
| Depression vs. No depression | 2014 | 0.000884 | -0.0004007 | 0.0021687 | 0.00042 | -0.0036702 | 0.0045102 |
| Depression vs. No depression | 2015 | 3.59E-06 | -0.0006752 | 0.0006824 | -0.0005892 | -0.0028773 | 0.001699 |
| Depression vs. No depression | 2017 | 0.0010077 | -0.0002425 | 0.0022579 | -0.0000375 | -0.0020361 | 0.0019611 |
| Depression vs. No depression | 2018 | -0.0001511 | -0.0011177 | 0.0008156 | -0.0002191 | -0.002422 | 0.0019839 |
| Depression vs. No depression | 2019 | 0.0004715 | -0.0007643 | 0.0017074 | 0.0000215 | -0.0021958 | 0.0022388 |
| Depression vs. No depression | 2020 | 0.0033126 | 0.0004628 | 0.0061624 | -0.0005685 | -0.0027642 | 0.0016273 |
| Depression vs. No depression | 2021 | -0.0017231 | -0.0039134 | 0.0004673 | 0.0015269 | -0.0014943 | 0.0045481 |
| Depression vs. No depression | 2022 | 0.0139398 | 0.0071739 | 0.0207057 | 0.0003895 | -0.0015063 | 0.0022852 |
| Depression vs. No depression | 2023 | 0.0120631 | 0.0006742 | 0.0234521 | -0.0010109 | -0.003307 | 0.0012853 |
| Diabetes vs. No diabetes | 2009 | 0.0000926 | -0.0002403 | 0.0004254 | -0.0001621 | -0.0010289 | 0.0007047 |
| Diabetes vs. No diabetes | 2010 | 0.0001227 | -0.0001998 | 0.0004453 | -0.0001138 | -0.0008691 | 0.0006414 |
| Diabetes vs. No diabetes | 2011 | 0.0000158 | -0.000163 | 0.0001945 | -0.0003692 | -0.0012547 | 0.0005163 |
| Diabetes vs. No diabetes | 2012 | 0.0001336 | -0.0002946 | 0.0005617 | -0.0000221 | -0.0008824 | 0.0008383 |
| Diabetes vs. No diabetes | 2013 | 0.0001322 | -0.0003869 | 0.0006513 | 0.0001862 | -0.0006352 | 0.0010076 |
| Diabetes vs. No diabetes | 2014 | 0.0000394 | -0.0003694 | 0.0004481 | 0.0000483 | -0.0072984 | 0.007395 |
| Diabetes vs. No diabetes | 2015 | 0.0000189 | -0.0006869 | 0.0007246 | 0.0004779 | -0.0002053 | 0.0011612 |
| Diabetes vs. No diabetes | 2017 | -0.0000127 | -0.0001353 | 0.00011 | -0.0002891 | -0.0011897 | 0.0006115 |
| Diabetes vs. No diabetes | 2019 | 5.81E-06 | -0.0000908 | 0.0001024 | -0.0005168 | -0.001492 | 0.0004584 |
| Diabetes vs. No diabetes | 2020 | -0.0000106 | -0.0001697 | 0.0001485 | -0.0002747 | -0.0012546 | 0.0007053 |
| Diabetes vs. No diabetes | 2021 | 0.0001065 | -0.0004808 | 0.0006937 | -0.0002925 | -0.0012566 | 0.0006716 |
| Diabetes vs. No diabetes | 2022 | 0.00083 | -0.0002879 | 0.0019478 | 0.0003794 | -0.0004926 | 0.0012514 |
| Diabetes vs. No diabetes | 2023 | 0.0009773 | -0.0008912 | 0.0028457 | 0.0005817 | -0.0005584 | 0.0017219 |
| Epilepsy vs. No epilepsy | 2009 | 0.0005767 | -0.0002566 | 0.00141 | 0.0004793 | -0.0004793 | 0.001438 |
| Epilepsy vs. No epilepsy | 2010 | 0.0005442 | -0.0001261 | 0.0012146 | 0.0006089 | -0.0002825 | 0.0015003 |
| Epilepsy vs. No epilepsy | 2011 | 0.0009523 | -0.0000846 | 0.0019892 | 0.0007026 | -0.0002525 | 0.0016577 |
| Epilepsy vs. No epilepsy | 2012 | 6.07E-06 | -0.0003383 | 0.0003505 | -0.0000865 | -0.0010472 | 0.0008742 |
| Epilepsy vs. No epilepsy | 2013 | 0.0000389 | -0.0002393 | 0.0003172 | 0.0000565 | -0.001086 | 0.0011989 |
| Epilepsy vs. No epilepsy | 2014 | -0.0000384 | -0.0002429 | 0.000166 | -0.0003614 | -0.2743166 | 0.2735939 |
| Epilepsy vs. No epilepsy | 2015 | 0.0007847 | -0.0000892 | 0.0016587 | 0.0007359 | -0.0000594 | 0.0015311 |
| Epilepsy vs. No epilepsy | 2017 | 0.0001394 | -0.0003558 | 0.0006346 | 0.0006062 | -0.0004239 | 0.0016363 |
| Epilepsy vs. No epilepsy | 2019 | 0.0000924 | -0.0002521 | 0.0004368 | -0.0003864 | -0.0016416 | 0.0008688 |
| Epilepsy vs. No epilepsy | 2020 | 0.0001567 | -0.0004783 | 0.0007917 | -0.0009844 | -0.0025758 | 0.000607 |
| Epilepsy vs. No epilepsy | 2021 | 0.0000564 | -0.0003621 | 0.0004749 | -0.0006232 | -0.0018534 | 0.000607 |
| Epilepsy vs. No epilepsy | 2022 | 2.12E-07 | -0.0001114 | 0.0001119 | -0.0002442 | -0.0013965 | 0.000908 |
| Epilepsy vs. No epilepsy | 2023 | -0.0000407 | -0.0004555 | 0.0003742 | 0.0003279 | -0.0011764 | 0.0018321 |
| Female vs. Male | 2009 | -0.0031077 | -0.0050617 | -0.0011537 | -0.0200033 | -0.0374418 | -0.0025648 |
| Female vs. Male | 2010 | -0.0004787 | -0.0013904 | 0.000433 | -0.0100748 | -0.0249939 | 0.0048443 |
| Female vs. Male | 2011 | 0.0002936 | -0.0007012 | 0.0012884 | -0.010379 | -0.0259273 | 0.0051693 |
| Female vs. Male | 2012 | 0.0008143 | -0.0003365 | 0.001965 | -0.0124789 | -0.0288472 | 0.0038894 |
| Female vs. Male | 2013 | 0.0011619 | -0.0000994 | 0.0024231 | -0.0161497 | -0.035842 | 0.0035425 |
| Female vs. Male | 2014 | 0.0005146 | -0.0002174 | 0.0012467 | -0.0003149 | -0.0147816 | 0.0141517 |
| Female vs. Male | 2015 | 0.0006571 | -0.0003538 | 0.0016679 | -0.0069775 | -0.0202368 | 0.0062818 |
| Female vs. Male | 2017 | 0.0000548 | -0.0014474 | 0.0015569 | -0.0277821 | -0.0721779 | 0.0166137 |
| Female vs. Male | 2019 | -0.0013485 | -0.0032168 | 0.0005198 | -0.0240815 | -0.0472325 | -0.0009306 |
| Female vs. Male | 2020 | -0.003086 | -0.0054861 | -0.0006859 | -0.0210038 | -0.0415031 | -0.0005045 |
| Female vs. Male | 2021 | -0.0033709 | -0.0062172 | -0.0005247 | -0.0320879 | -0.0559159 | -0.00826 |
| Female vs. Male | 2022 | -0.0018333 | -0.0037771 | 0.0001104 | -0.0089416 | -0.0294076 | 0.0115245 |
| Female vs. Male | 2023 | -0.0015763 | -0.0041269 | 0.0009743 | -0.0140547 | -0.0407972 | 0.0126878 |
| GCSE maximum vs. Above GCSE | 2009 | 0.0092041 | 0.0059829 | 0.0124253 | 0.0047563 | -0.006299 | 0.0158117 |
| GCSE maximum vs. Above GCSE | 2010 | 0.0059538 | 0.00388 | 0.0080276 | 0.006897 | -0.0046147 | 0.0184087 |
| GCSE maximum vs. Above GCSE | 2011 | 0.0028232 | 0.0011808 | 0.0044656 | -0.0007231 | -0.010515 | 0.0090688 |
| GCSE maximum vs. Above GCSE | 2012 | 0.001955 | 0.0004446 | 0.0034653 | -0.0004469 | -0.0103962 | 0.0095025 |
| GCSE maximum vs. Above GCSE | 2013 | 0.0009459 | -0.0000949 | 0.0019866 | -0.00491 | -0.0133514 | 0.0035313 |
| GCSE maximum vs. Above GCSE | 2014 | -7.55E-06 | -0.0012948 | 0.0012797 | 0.0009322 | -0.0055079 | 0.0073724 |
| GCSE maximum vs. Above GCSE | 2015 | -8.35E-06 | -0.0010331 | 0.0010164 | -0.0038067 | -0.0276519 | 0.0200385 |
| GCSE maximum vs. Above GCSE | 2017 | 0.0009389 | -0.0003225 | 0.0022004 | -0.0023737 | -0.0111281 | 0.0063807 |
| GCSE maximum vs. Above GCSE | 2019 | 0.0014737 | -0.0001528 | 0.0031002 | -0.0014192 | -0.0126279 | 0.0097896 |
| GCSE maximum vs. Above GCSE | 2020 | 0.0019937 | -0.0001073 | 0.0040947 | 0.0010125 | -0.0114938 | 0.0135189 |
| GCSE maximum vs. Above GCSE | 2021 | 0.0034934 | 0.0006668 | 0.0063201 | 0.0072336 | -0.0069419 | 0.0214092 |
| GCSE maximum vs. Above GCSE | 2022 | 0.0055241 | 0.0021652 | 0.0088831 | 0.0035401 | -0.010359 | 0.0174392 |
| GCSE maximum vs. Above GCSE | 2023 | 0.0056411 | 0.0003564 | 0.0109259 | -0.0022166 | -0.0186009 | 0.0141677 |
| Has condition vs. No condition | 2009 | -0.0019546 | -0.003876 | -0.0000331 | -0.0348044 | -0.0764089 | 0.0068001 |
| Has condition vs. No condition | 2010 | -0.0018505 | -0.0033543 | -0.0003466 | -0.0308346 | -0.0609938 | -0.0006753 |
| Has condition vs. No condition | 2011 | -0.0021839 | -0.0037926 | -0.0005752 | -0.0404668 | -0.0750517 | -0.0058819 |
| Has condition vs. No condition | 2012 | -0.0018041 | -0.0034223 | -0.000186 | -0.037581 | -0.0722566 | -0.0029053 |
| Has condition vs. No condition | 2013 | -0.0016805 | -0.0031992 | -0.0001619 | -0.0352154 | -0.0639411 | -0.0064897 |
| Has condition vs. No condition | 2014 | -0.0034512 | -0.0055321 | -0.0013703 | -0.0053876 | -0.0323901 | 0.0216149 |
| Has condition vs. No condition | 2015 | -0.0009831 | -0.0029003 | 0.0009341 | -0.0052493 | -0.1194966 | 0.1089979 |
| Has condition vs. No condition | 2017 | 0.0025769 | 0.0003741 | 0.0047797 | -0.0074846 | -0.0409521 | 0.025983 |
| Has condition vs. No condition | 2018 | 0.0027704 | 0.0007439 | 0.0047968 | -0.0194912 | -0.0647323 | 0.0257498 |
| Has condition vs. No condition | 2019 | 0.0058761 | 0.0027834 | 0.0089688 | 0.0004243 | -0.046004 | 0.0468525 |
| Has condition vs. No condition | 2020 | 0.0061303 | 0.0028548 | 0.0094059 | -0.0247285 | -0.0697844 | 0.0203274 |
| Has condition vs. No condition | 2021 | 0.0029609 | 0.0003637 | 0.005558 | -0.0313869 | -0.0776828 | 0.014909 |
| Has condition vs. No condition | 2022 | 0.0081138 | 0.0040187 | 0.0122088 | 0.0336555 | -0.0458193 | 0.1131304 |
| Has condition vs. No condition | 2023 | 0.0100166 | 0.0038144 | 0.0162188 | -0.0071037 | -0.0718302 | 0.0576229 |
| High blood pressure vs. normal | 2009 | 0.0000135 | -0.001099 | 0.001126 | -0.0010456 | -0.0022827 | 0.0001915 |
| High blood pressure vs. normal | 2010 | 0.0002655 | -0.0007611 | 0.0012922 | -0.0008554 | -0.0019526 | 0.0002417 |
| High blood pressure vs. normal | 2011 | 0.0012906 | -0.0001114 | 0.0026925 | -0.0003737 | -0.0014845 | 0.0007371 |
| High blood pressure vs. normal | 2012 | 0.0009388 | -0.0003063 | 0.0021839 | -0.000393 | -0.0015571 | 0.000771 |
| High blood pressure vs. normal | 2013 | 0.0002436 | -0.0003997 | 0.0008869 | -0.0006016 | -0.0017919 | 0.0005888 |
| High blood pressure vs. normal | 2014 | -0.0001781 | -0.0006432 | 0.0002869 | -0.0003404 | -0.0263916 | 0.0257108 |
| High blood pressure vs. normal | 2015 | 0.0002037 | -0.0003259 | 0.0007333 | -0.0002893 | -0.0016081 | 0.0010294 |
| High blood pressure vs. normal | 2017 | -0.0004331 | -0.001292 | 0.0004258 | 0.0002705 | -0.0010694 | 0.0016103 |
| High blood pressure vs. normal | 2019 | -0.0005162 | -0.0018022 | 0.0007698 | -0.0001957 | -0.0019395 | 0.001548 |
| High blood pressure vs. normal | 2020 | -0.0001562 | -0.0006785 | 0.000366 | -0.0004508 | -0.0019213 | 0.0010197 |
| High blood pressure vs. normal | 2021 | -0.0016376 | -0.0035329 | 0.0002576 | 0.0010537 | -0.0008139 | 0.0029213 |
| High blood pressure vs. normal | 2022 | 0.0000724 | -0.0012462 | 0.0013909 | 0.0006893 | -0.0008527 | 0.0022313 |
| High blood pressure vs. normal | 2023 | -0.0000281 | -0.0005408 | 0.0004845 | -0.0015469 | -0.0034682 | 0.0003743 |
| London & South vs. Other | 2009 | -0.0008245 | -0.0019745 | 0.0003254 | -0.0096227 | -0.022146 | 0.0029005 |
| London & South vs. Other | 2010 | -0.000726 | -0.0015727 | 0.0001208 | -0.0080958 | -0.019332 | 0.0031405 |
| London & South vs. Other | 2011 | -0.0001257 | -0.0005068 | 0.0002554 | -0.0008562 | -0.0119877 | 0.0102754 |
| London & South vs. Other | 2012 | -0.000017 | -0.0002785 | 0.0002446 | 0.0003052 | -0.0111055 | 0.0117159 |
| London & South vs. Other | 2013 | 0.0000188 | -0.000126 | 0.0001636 | 0.0020919 | -0.0091569 | 0.0133408 |
| London & South vs. Other | 2014 | -0.0000769 | -0.000448 | 0.0002942 | -0.0012925 | -0.0115149 | 0.0089299 |
| London & South vs. Other | 2015 | 0.0000529 | -0.000295 | 0.0004008 | -0.0010494 | -0.0119929 | 0.0098941 |
| London & South vs. Other | 2017 | 0.0000192 | -0.0002188 | 0.0002573 | 0.0006553 | -0.0115232 | 0.0128338 |
| London & South vs. Other | 2019 | 0.0000714 | -0.0006027 | 0.0007455 | 0.0047564 | -0.0080129 | 0.0175257 |
| London & South vs. Other | 2020 | -0.0003534 | -0.0011734 | 0.0004666 | -0.0010287 | -0.0146807 | 0.0126232 |
| London & South vs. Other | 2021 | -0.0002259 | -0.0011116 | 0.0006597 | 0.001094 | -0.0130816 | 0.0152696 |
| London & South vs. Other | 2022 | -0.000472 | -0.0016376 | 0.0006937 | -0.0009153 | -0.015028 | 0.0131974 |
| London & South vs. Other | 2023 | -0.0004746 | -0.0020645 | 0.0011154 | -0.0008143 | -0.0190617 | 0.017433 |
| Lowest quintile vs. Other quintiles | 2009 | 0.0050638 | 0.0022844 | 0.0078432 | 0.0018777 | -0.0047128 | 0.0084682 |
| Lowest quintile vs. Other quintiles | 2010 | 0.0033311 | 0.0013383 | 0.0053238 | 0.0010504 | -0.004827 | 0.0069277 |
| Lowest quintile vs. Other quintiles | 2011 | 0.0015335 | -0.0007351 | 0.003802 | 0.0030007 | -0.0036731 | 0.0096745 |
| Lowest quintile vs. Other quintiles | 2012 | 0.0013382 | -0.0009465 | 0.0036228 | 0.0028729 | -0.0040555 | 0.0098013 |
| Lowest quintile vs. Other quintiles | 2013 | 0.0008485 | -0.0010044 | 0.0027014 | -0.0006985 | -0.006201 | 0.0048041 |
| Lowest quintile vs. Other quintiles | 2014 | 0.000745 | -0.0012979 | 0.0027879 | 0.0012647 | -0.0035915 | 0.0061209 |
| Lowest quintile vs. Other quintiles | 2015 | -0.0024647 | -0.0046935 | -0.0002358 | -0.0007533 | -0.0381822 | 0.0366757 |
| Lowest quintile vs. Other quintiles | 2017 | 0.0013547 | -0.0008589 | 0.0035684 | 0.0010315 | -0.0073045 | 0.0093676 |
| Lowest quintile vs. Other quintiles | 2019 | 0.0021857 | -0.0007119 | 0.0050834 | 0.0045386 | -0.0055224 | 0.0145996 |
| Lowest quintile vs. Other quintiles | 2020 | 0.0031854 | 0.0002746 | 0.0060961 | -0.0007053 | -0.0080774 | 0.0066668 |
| Lowest quintile vs. Other quintiles | 2021 | 0.0043355 | 0.0011877 | 0.0074832 | -0.0019848 | -0.0092597 | 0.0052901 |
| Lowest quintile vs. Other quintiles | 2022 | 0.0062851 | 0.0023701 | 0.0102002 | 0.0047185 | -0.0064388 | 0.0158758 |
| Lowest quintile vs. Other quintiles | 2023 | 0.0025042 | -0.0012643 | 0.0062728 | -0.0037413 | -0.0136343 | 0.0061516 |
| Most deprived vs. Less deprived | 2009 | 0.0062506 | 0.0036597 | 0.0088415 | -0.0002736 | -0.0079387 | 0.0073914 |
| Most deprived vs. Less deprived | 2010 | 0.0049875 | 0.00311 | 0.0068651 | 0.0028047 | -0.0050218 | 0.0106312 |
| Most deprived vs. Less deprived | 2011 | 0.0020115 | 0.0001257 | 0.0038972 | 0.0041575 | -0.0040514 | 0.0123664 |
| Most deprived vs. Less deprived | 2012 | 0.0017265 | -0.0002982 | 0.0037512 | 0.0058135 | -0.0033328 | 0.0149598 |
| Most deprived vs. Less deprived | 2013 | 0.000959 | -0.00072 | 0.0026379 | 0.0026259 | -0.0062322 | 0.011484 |
| Most deprived vs. Less deprived | 2014 | -0.0001457 | -0.0017791 | 0.0014877 | 0.0016332 | -0.0064022 | 0.0096686 |
| Most deprived vs. Less deprived | 2015 | -0.0006886 | -0.0020533 | 0.0006761 | -0.0004901 | -0.0099967 | 0.0090164 |
| Most deprived vs. Less deprived | 2017 | -0.0002873 | -0.0014148 | 0.0008401 | -0.0034224 | -0.0095688 | 0.002724 |
| Most deprived vs. Less deprived | 2019 | -0.0003187 | -0.0015215 | 0.0008841 | -0.0044583 | -0.012166 | 0.0032494 |
| Most deprived vs. Less deprived | 2020 | -0.0015596 | -0.0036411 | 0.0005219 | -0.0004155 | -0.0100624 | 0.0092315 |
| Most deprived vs. Less deprived | 2021 | 0.0000334 | -0.0015402 | 0.0016069 | -0.0037495 | -0.0130809 | 0.0055819 |
| Most deprived vs. Less deprived | 2022 | -0.0003683 | -0.0024537 | 0.001717 | 0.0014674 | -0.0086172 | 0.0115519 |
| Most deprived vs. Less deprived | 2023 | 0.0029496 | -0.0009096 | 0.0068089 | 0.0030565 | -0.0098459 | 0.0159589 |
| Non white vs. White | 2009 | 0.0015718 | -0.0019642 | 0.0051077 | 0.000471 | -0.0054653 | 0.0064074 |
| Non white vs. White | 2010 | -0.000654 | -0.0022148 | 0.0009067 | -0.0019492 | -0.0072302 | 0.0033318 |
| Non white vs. White | 2011 | -0.000128 | -0.0004665 | 0.0002105 | -0.0037325 | -0.0094596 | 0.0019945 |
| Non white vs. White | 2012 | -0.0000243 | -0.000157 | 0.0001084 | -0.0002029 | -0.00588 | 0.0054742 |
| Non white vs. White | 2013 | 8.23E-06 | -0.0001266 | 0.000143 | -0.0007126 | -0.0063199 | 0.0048946 |
| Non white vs. White | 2014 | -1.27E-07 | -0.0000969 | 0.0000967 | -0.0017559 | -0.0074954 | 0.0039835 |
| Non white vs. White | 2015 | 0.0001203 | -0.0005068 | 0.0007474 | -0.001605 | -0.0074778 | 0.0042678 |
| Non white vs. White | 2017 | -0.0000894 | -0.0004257 | 0.0002469 | -0.002296 | -0.0087954 | 0.0042035 |
| Non white vs. White | 2019 | -0.0004615 | -0.0015372 | 0.0006141 | -0.0027648 | -0.0091071 | 0.0035776 |
| Non white vs. White | 2020 | 0.0007654 | -0.0008302 | 0.0023611 | 0.0013821 | -0.0054985 | 0.0082626 |
| Non white vs. White | 2021 | -0.0006603 | -0.0031677 | 0.001847 | -0.0022323 | -0.0091248 | 0.0046603 |
| Non white vs. White | 2022 | -0.0017471 | -0.004227 | 0.0007329 | -0.0044587 | -0.0115061 | 0.0025887 |
| Non white vs. White | 2023 | -0.0019907 | -0.0091048 | 0.0051234 | -0.0029074 | -0.0114958 | 0.005681 |
| Psychological distress vs. No distress | 2009 | -0.0016519 | -0.0032529 | -0.0000509 | -0.0049437 | -0.0124733 | 0.002586 |
| Psychological distress vs. No distress | 2010 | -0.0019915 | -0.0033695 | -0.0006135 | -0.0039706 | -0.0099331 | 0.0019919 |
| Psychological distress vs. No distress | 2011 | -0.0018779 | -0.0033004 | -0.0004554 | -0.0043928 | -0.0108433 | 0.0020577 |
| Psychological distress vs. No distress | 2012 | 0.0000379 | -0.0014174 | 0.0014931 | -0.0021735 | -0.0088541 | 0.0045071 |
| Psychological distress vs. No distress | 2013 | -0.0001447 | -0.0009526 | 0.0006631 | -0.0066595 | -0.0117811 | -0.0015379 |
| Psychological distress vs. No distress | 2014 | -0.0000398 | -0.0007731 | 0.0006934 | -0.040213 | -0.7976632 | 0.7172372 |
| Psychological distress vs. No distress | 2015 | -0.0006539 | -0.001912 | 0.0006042 | -0.0036529 | -0.0159588 | 0.0086529 |
| Psychological distress vs. No distress | 2017 | 0.0017247 | -9.56E-06 | 0.0034589 | -0.0011415 | -0.0075135 | 0.0052305 |
| Psychological distress vs. No distress | 2018 | 0.0021696 | 0.0002902 | 0.004049 | -0.0045301 | -0.0138944 | 0.0048342 |
| Psychological distress vs. No distress | 2019 | 0.0042943 | 0.0017054 | 0.0068831 | -0.0012145 | -0.0087374 | 0.0063084 |
| Psychological distress vs. No distress | 2020 | 0.0071403 | 0.0033475 | 0.0109332 | -0.003538 | -0.0115046 | 0.0044286 |
| Psychological distress vs. No distress | 2021 | 0.0071376 | 0.0031137 | 0.0111614 | -0.0023009 | -0.0105802 | 0.0059784 |
| Psychological distress vs. No distress | 2022 | 0.0075799 | 0.0036257 | 0.011534 | 0.0009386 | -0.0083531 | 0.0102304 |
| Psychological distress vs. No distress | 2023 | 0.0042005 | -0.0001375 | 0.0085384 | -0.0058169 | -0.0170774 | 0.0054436 |

### Supplementary file S8. Fixed effects Poisson regression estimates with 2016 as year reference

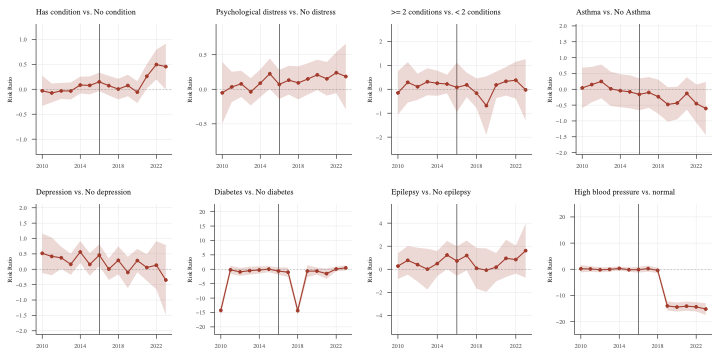

|  | Variable | Est | Cim | Cip |
| --- | --- | --- | --- | --- |
| 2010 | Has condition vs. No condition | -0.0302795 | -0.332923 | 0.272364 |
| 2011 | Has condition vs. No condition | -0.0743232 | -0.2633509 | 0.1147046 |
| 2012 | Has condition vs. No condition | -0.0322989 | -0.1918264 | 0.1272287 |
| 2013 | Has condition vs. No condition | -0.0344222 | -0.2052492 | 0.1364048 |
| 2014 | Has condition vs. No condition | 0.0846796 | -0.0831818 | 0.252541 |
| 2015 | Has condition vs. No condition | 0.0775391 | -0.10244 | 0.2575182 |
| 2016 | Has condition vs. No condition | 0.1489547 | -0.0354095 | 0.3333189 |
| 2017 | Has condition vs. No condition | 0.0723233 | -0.1241345 | 0.2687811 |
| 2018 | Has condition vs. No condition | 0.0038723 | -0.2027881 | 0.2105326 |
| 2019 | Has condition vs. No condition | 0.0754817 | -0.1393142 | 0.2902776 |
| 2020 | Has condition vs. No condition | -0.0578389 | -0.2768094 | 0.1611316 |
| 2021 | Has condition vs. No condition | 0.2593891 | 0.014896 | 0.5038822 |
| 2022 | Has condition vs. No condition | 0.4952504 | 0.1962727 | 0.7942282 |
| 2023 | Has condition vs. No condition | 0.4536522 | -0.0076324 | 0.9149368 |
| 2010 | Psychological distress vs. No distress | -0.051401 | -0.4973563 | 0.3945542 |
| 2011 | Psychological distress vs. No distress | 0.033819 | -0.1856941 | 0.253332 |
| 2012 | Psychological distress vs. No distress | 0.0785951 | -0.110331 | 0.2675212 |
| 2013 | Psychological distress vs. No distress | -0.0373132 | -0.2343552 | 0.1597288 |
| 2014 | Psychological distress vs. No distress | 0.0874653 | -0.1120846 | 0.2870152 |
| 2015 | Psychological distress vs. No distress | 0.2245985 | 0.0042749 | 0.4449221 |
| 2016 | Psychological distress vs. No distress | 0.0702915 | -0.1387184 | 0.2793013 |
| 2017 | Psychological distress vs. No distress | 0.1315995 | -0.0772437 | 0.3404428 |
| 2018 | Psychological distress vs. No distress | 0.0925865 | -0.1464748 | 0.3316478 |
| 2019 | Psychological distress vs. No distress | 0.1492631 | -0.0769877 | 0.3755138 |
| 2020 | Psychological distress vs. No distress | 0.2085748 | -0.0117166 | 0.4288663 |
| 2021 | Psychological distress vs. No distress | 0.1512478 | -0.0914085 | 0.393904 |
| 2022 | Psychological distress vs. No distress | 0.2393502 | -0.0564982 | 0.5351986 |
| 2023 | Psychological distress vs. No distress | 0.1845647 | -0.2913857 | 0.660515 |
| 2010 | >= 2 conditions vs. < 2 conditions | -0.1498959 | -1.053635 | 0.7538433 |
| 2011 | >= 2 conditions vs. < 2 conditions | 0.2899461 | -0.5555439 | 1.135436 |
| 2012 | >= 2 conditions vs. < 2 conditions | 0.1045058 | -0.4414697 | 0.6504813 |
| 2013 | >= 2 conditions vs. < 2 conditions | 0.3137158 | -0.2486427 | 0.8760743 |
| 2014 | >= 2 conditions vs. < 2 conditions | 0.2520046 | -0.2732239 | 0.7772331 |
| 2015 | >= 2 conditions vs. < 2 conditions | 0.2189741 | -0.1727507 | 0.6106988 |
| 2016 | >= 2 conditions vs. < 2 conditions | 0.0786071 | -0.9534547 | 1.110669 |
| 2017 | >= 2 conditions vs. < 2 conditions | 0.1831803 | -0.3282131 | 0.6945737 |
| 2018 | >= 2 conditions vs. < 2 conditions | -0.1631719 | -0.7801216 | 0.4537777 |
| 2019 | >= 2 conditions vs. < 2 conditions | -0.6845422 | -1.907678 | 0.5385932 |
| 2020 | >= 2 conditions vs. < 2 conditions | 0.1801929 | -0.3745745 | 0.7349604 |
| 2021 | >= 2 conditions vs. < 2 conditions | 0.3324641 | -0.2646364 | 0.9295647 |
| 2022 | >= 2 conditions vs. < 2 conditions | 0.3775212 | -0.3937161 | 1.148759 |
| 2023 | >= 2 conditions vs. < 2 conditions | -0.0253791 | -1.307109 | 1.256351 |
| 2010 | Asthma vs. No Asthma | 0.0426285 | -0.5956763 | 0.6809333 |
| 2011 | Asthma vs. No Asthma | 0.1480568 | -0.4117802 | 0.7078937 |
| 2012 | Asthma vs. No Asthma | 0.2466991 | -0.2875458 | 0.780944 |
| 2013 | Asthma vs. No Asthma | 0.0136581 | -0.5284353 | 0.5557515 |
| 2014 | Asthma vs. No Asthma | -0.0446676 | -0.5638318 | 0.4744966 |
| 2015 | Asthma vs. No Asthma | -0.0808014 | -0.6027626 | 0.4411598 |
| 2016 | Asthma vs. No Asthma | -0.1608161 | -0.664288 | 0.3426558 |
| 2017 | Asthma vs. No Asthma | -0.1007253 | -0.5863375 | 0.3848869 |
| 2018 | Asthma vs. No Asthma | -0.2351427 | -0.7747585 | 0.3044731 |
| 2019 | Asthma vs. No Asthma | -0.4792051 | -1.022964 | 0.0645541 |
| 2020 | Asthma vs. No Asthma | -0.4337071 | -0.9469297 | 0.0795155 |
| 2021 | Asthma vs. No Asthma | -0.1313734 | -0.6427478 | 0.3800011 |
| 2022 | Asthma vs. No Asthma | -0.4518488 | -1.055 | 0.1513028 |
| 2023 | Asthma vs. No Asthma | -0.6064471 | -1.441483 | 0.2285885 |
| 2010 | Depression vs. No depression | 0.5187348 | -0.1231389 | 1.160608 |
| 2011 | Depression vs. No depression | 0.4190535 | -0.1930301 | 1.031137 |
| 2012 | Depression vs. No depression | 0.3739367 | 0.0149297 | 0.7329438 |
| 2013 | Depression vs. No depression | 0.1617349 | -0.1816043 | 0.5050742 |
| 2014 | Depression vs. No depression | 0.5620049 | 0.2042679 | 0.919742 |
| 2015 | Depression vs. No depression | 0.1574097 | -0.2084838 | 0.5233033 |
| 2016 | Depression vs. No depression | 0.460089 | 0.1033978 | 0.8167802 |
| 2017 | Depression vs. No depression | 0.0046016 | -0.3453931 | 0.3545962 |
| 2018 | Depression vs. No depression | 0.2902471 | -0.165326 | 0.7458201 |
| 2019 | Depression vs. No depression | -0.1034702 | -0.6135713 | 0.4066308 |
| 2020 | Depression vs. No depression | 0.2830874 | -0.0946593 | 0.660834 |
| 2021 | Depression vs. No depression | 0.0567932 | -0.3742278 | 0.4878141 |
| 2022 | Depression vs. No depression | 0.1324215 | -0.6494294 | 0.9142725 |
| 2023 | Depression vs. No depression | -0.3491943 | -1.484946 | 0.7865575 |
| 2010 | Diabetes vs. No diabetes | -14.28283 | -15.84889 | -12.71676 |
| 2011 | Diabetes vs. No diabetes | -0.1999072 | -1.61493 | 1.215116 |
| 2012 | Diabetes vs. No diabetes | -0.9055548 | -2.284392 | 0.4732828 |
| 2013 | Diabetes vs. No diabetes | -0.4796238 | -1.861417 | 0.9021692 |
| 2014 | Diabetes vs. No diabetes | -0.2410849 | -1.509961 | 1.027792 |
| 2015 | Diabetes vs. No diabetes | 0.0341586 | -1.014399 | 1.082716 |
| 2016 | Diabetes vs. No diabetes | -0.6159124 | -1.899657 | 0.6678321 |
| 2017 | Diabetes vs. No diabetes | -1.014467 | -2.630769 | 0.6018345 |
| 2018 | Diabetes vs. No diabetes | -14.36625 | -15.69059 | -13.04191 |
| 2019 | Diabetes vs. No diabetes | -0.6277737 | -2.682806 | 1.427258 |
| 2020 | Diabetes vs. No diabetes | -0.6300383 | -1.847319 | 0.5872428 |
| 2021 | Diabetes vs. No diabetes | -1.522508 | -3.315935 | 0.2709201 |
| 2022 | Diabetes vs. No diabetes | 0.0851569 | -0.7127198 | 0.8830335 |
| 2023 | Diabetes vs. No diabetes | 0.4711065 | -0.4564797 | 1.398693 |
| 2010 | Epilepsy vs. No epilepsy | 0.2743385 | -0.8410965 | 1.389773 |
| 2011 | Epilepsy vs. No epilepsy | 0.7750271 | -0.4701861 | 2.02024 |
| 2012 | Epilepsy vs. No epilepsy | 0.400483 | -1.063304 | 1.86427 |
| 2013 | Epilepsy vs. No epilepsy | 0.0065101 | -1.760827 | 1.773847 |
| 2014 | Epilepsy vs. No epilepsy | 0.4932336 | -0.5995687 | 1.586036 |
| 2015 | Epilepsy vs. No epilepsy | 1.236372 | 0.0128437 | 2.459901 |
| 2016 | Epilepsy vs. No epilepsy | 0.716913 | -0.5570746 | 1.990901 |
| 2017 | Epilepsy vs. No epilepsy | 1.193512 | -0.1013768 | 2.488401 |
| 2018 | Epilepsy vs. No epilepsy | 0.0843591 | -1.68162 | 1.850338 |
| 2019 | Epilepsy vs. No epilepsy | -0.0763636 | -1.964731 | 1.812003 |
| 2020 | Epilepsy vs. No epilepsy | 0.1765813 | -1.050833 | 1.403996 |
| 2021 | Epilepsy vs. No epilepsy | 0.9484164 | -0.6683753 | 2.565208 |
| 2022 | Epilepsy vs. No epilepsy | 0.8410608 | -0.3733433 | 2.055465 |
| 2023 | Epilepsy vs. No epilepsy | 1.614011 | -0.7321576 | 3.96018 |
| 2010 | High blood pressure vs. normal | 0.1921653 | -1.208391 | 1.592722 |
| 2011 | High blood pressure vs. normal | 0.1147158 | -0.9407058 | 1.170137 |
| 2012 | High blood pressure vs. normal | -0.1859959 | -1.164723 | 0.7927315 |
| 2013 | High blood pressure vs. normal | -0.0466084 | -0.9131117 | 0.819895 |
| 2014 | High blood pressure vs. normal | 0.3085887 | -0.5134419 | 1.130619 |
| 2015 | High blood pressure vs. normal | -0.1671127 | -0.9771668 | 0.6429414 |
| 2016 | High blood pressure vs. normal | -0.1591434 | -1.286086 | 0.9677989 |
| 2017 | High blood pressure vs. normal | 0.2383229 | -0.9102426 | 1.386888 |
| 2018 | High blood pressure vs. normal | -0.4194632 | -1.464338 | 0.6254116 |
| 2019 | High blood pressure vs. normal | -14.01272 | -15.73983 | -12.28561 |
| 2020 | High blood pressure vs. normal | -14.44678 | -16.16933 | -12.72423 |
| 2021 | High blood pressure vs. normal | -14.103 | -15.87197 | -12.33403 |
| 2022 | High blood pressure vs. normal | -14.36479 | -16.13594 | -12.59365 |
| 2023 | High blood pressure vs. normal | -15.20617 | -17.45306 | -12.95929 |

### Supplementary file S9. Multivariate models including self-report health condition or disability and GHQ-12 caseness

|  | **2009-13 to 2014-18** | | |  | **2019-23 to 2014-18** | | |  | FE  main effect  (2014-18) |
| --- | --- | --- | --- | --- | --- | --- | --- | --- | --- |
|  | Decomposition | | FE  interaction |  | Decomposition | | FE  interaction |  |  |
|  | Explained component | Unexplained component |  |  | Explained component | Unexplained component |  |  |  |
| Self-reported health conditions or disability | -0.178^***^ | -2.301^**^ | -0.073 |  | 0.494^***^ | 0.431 | 0.065 |  | 0.061 |
|  | (-0.250;-0.107) | (-4.142;-0.459) | (-0.244;0.096) |  | (0.345;0.642) | (-2.12; 2.98) | (-0.116;0.248) |  | (-0.062;0.185) |
| Psychological distress (GHQ-12 caseness) | -0.162^***^ | -0.057 | -0.086 |  | 0.400^***^ | -0.160 | 0.055 |  | 0.125^**^ |
|  | (-0.224;-0.101) | (-0.282;0.398) | (-0.246;0.074) |  | (0.266;0.533) | (-0.289;0.610) | (-0.110;0.222) |  | (0.017;0.233) |

### Supplementary file S10. Blinder-Oaxaca Decomposition (probit) stratification

#### Blinder Oaxaca Decomposition (probit), stratification by sex

Male

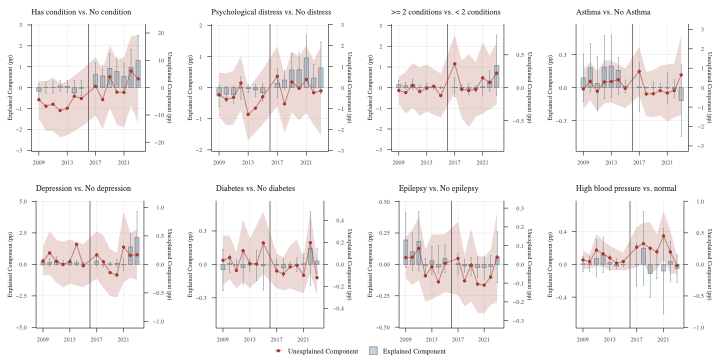

Female

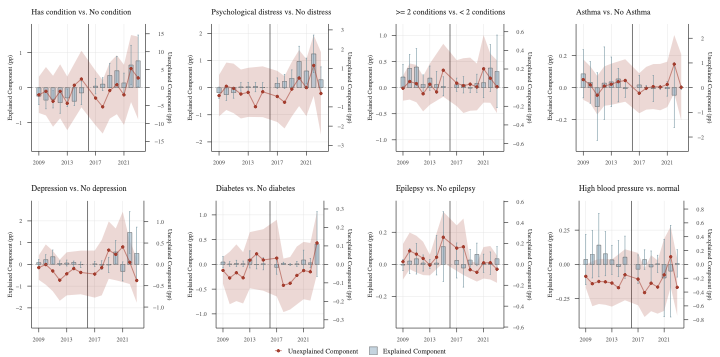

#### Blinder-Oaxaca Decomposition (probit), stratification by age group

16-19

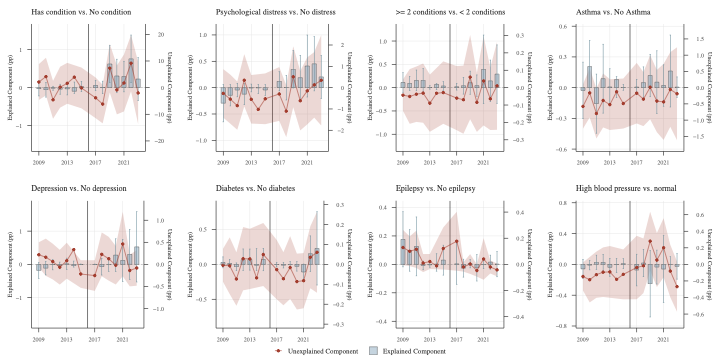

20-24

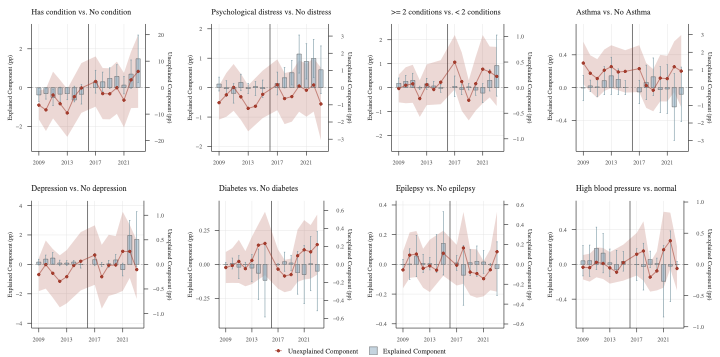

#### Blinder-Oaxaca Decomposition (probit), stratification by ethnicity

Non-white

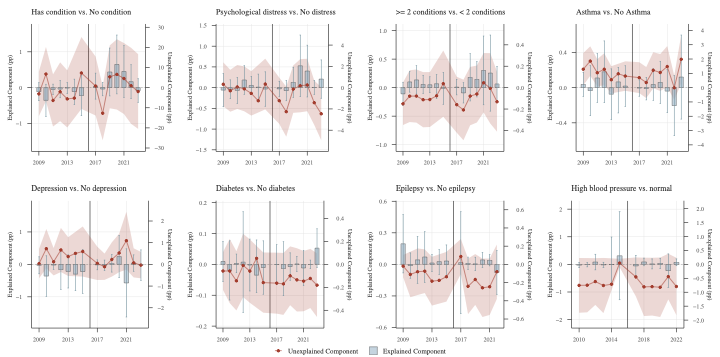

White

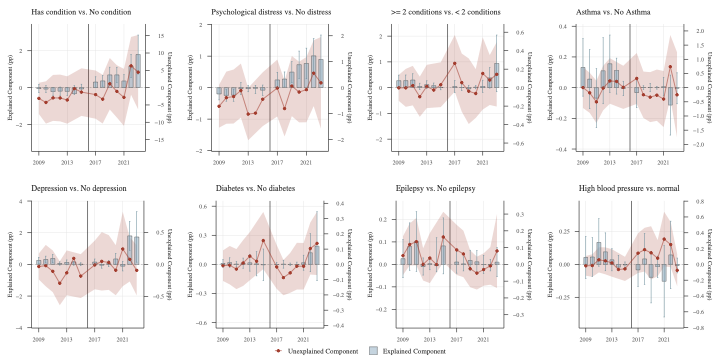

#### Blinder-Oaxaca Decomposition (probit), stratification by IMD

Most deprived

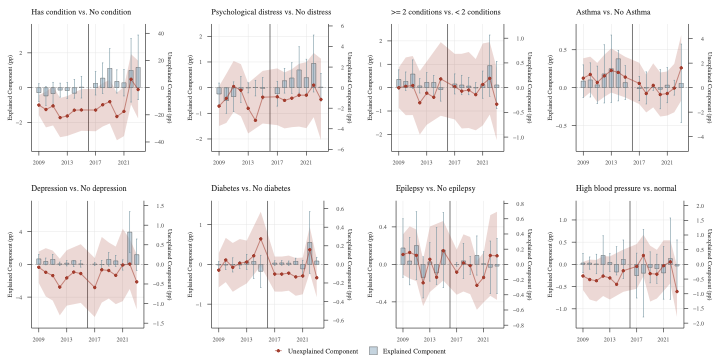

Not most deprived

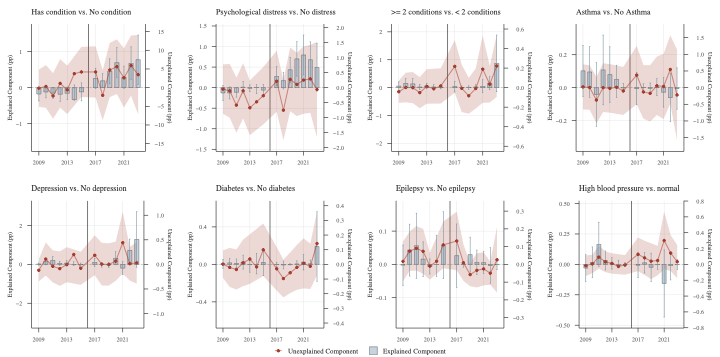

#### Blinder-Oaxaca Decomposition (probit), stratification by child.ren responsibilities

Children responsibilities

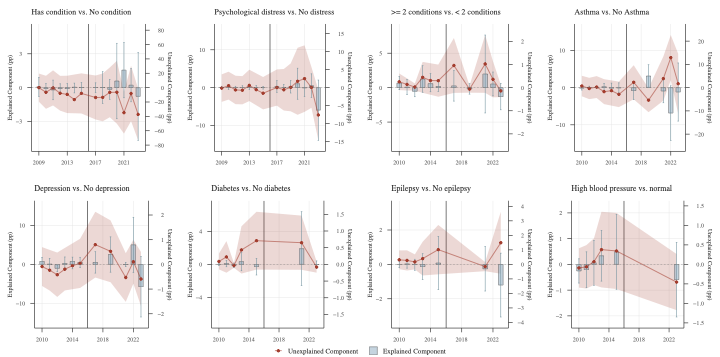

No children responsibility

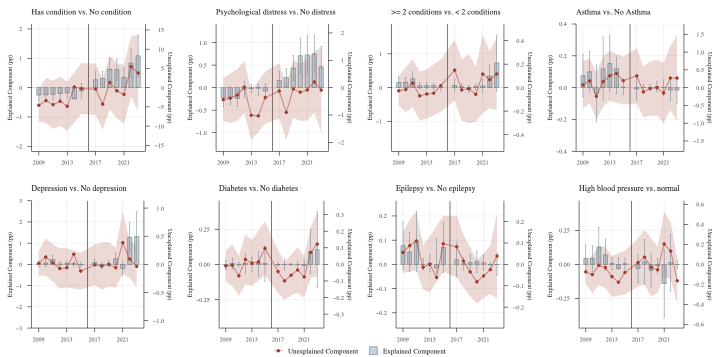

#### Blinder-Oaxaca Decomposition (probit), stratification by education

GCSE or below

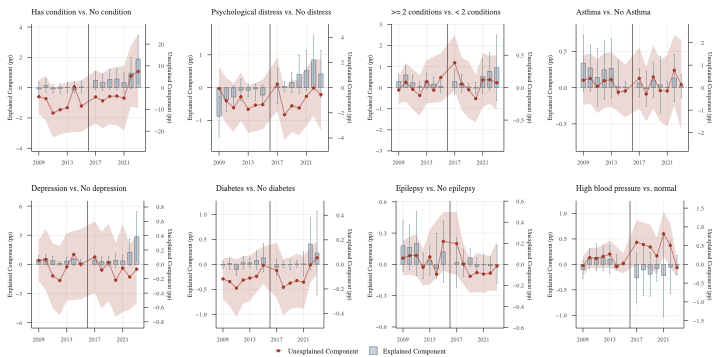

Above GCSE

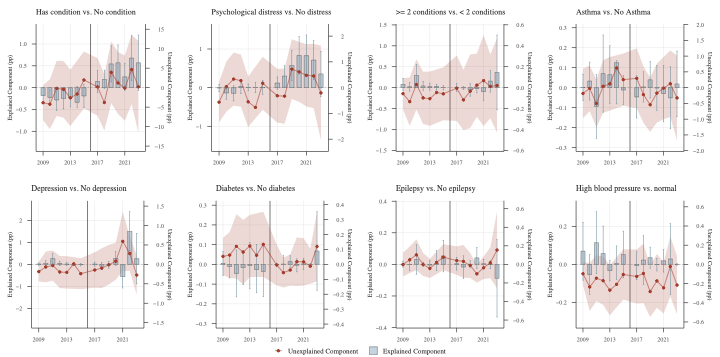

#### Blinder-Oaxaca Decomposition (probit), stratification by household incomes

Lowest quantile

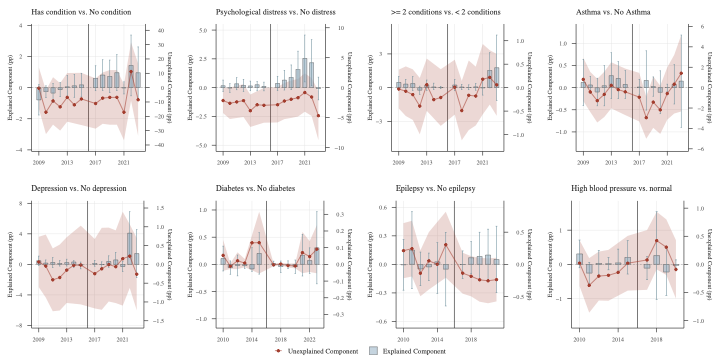

Not lowest quantile
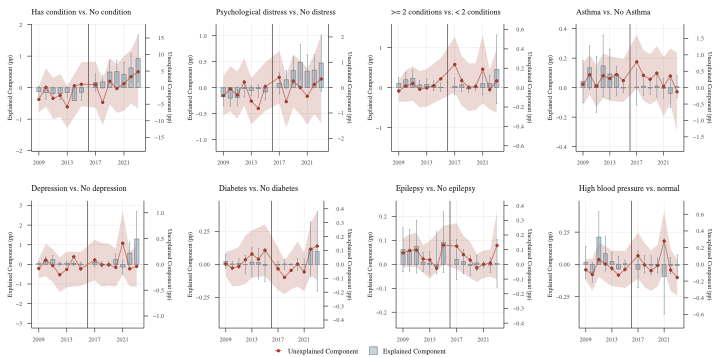

#### Blinder-Oaxaca Decomposition (probit), stratification by region

London & South of England

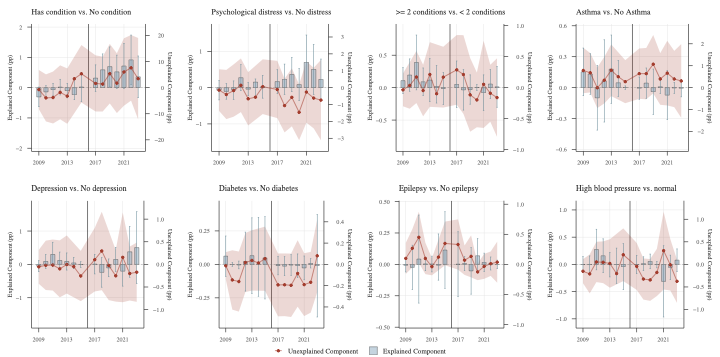

Not London & South of England

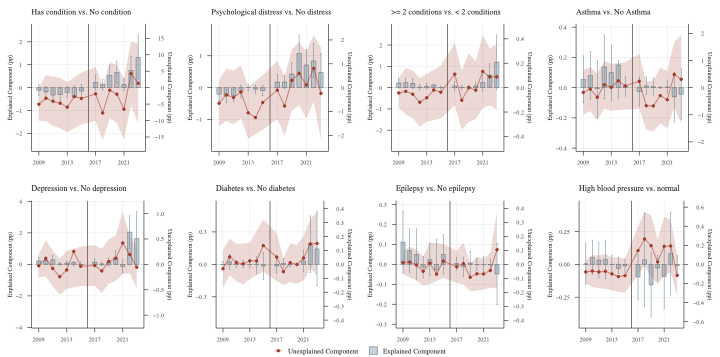

### Supplementary file S11. Fixed effect Poisson Regression by sex

|  | **Health** | | **Psychological distress** | | **Multimorbidity** | | **Asthma** | |
| --- | --- | --- | --- | --- | --- | --- | --- | --- |
|  | Female | Male | Female | Male | Female | Male | Female | Male |
| Period 2009-2013 | -0.096 | 0.035 | -0.095 | 0.067 | -0.151 | 0.058 | -0.232^**^ | 0.046 |
|  | [-0.230,0.038] | [-0.078,0.148] | [-0.249,0.058] | [-0.064,0.199] | [-0.316,0.013] | [-0.077,0.195] | [-0.406,-0.058] | [-0.110,0.203] |
| Period 2019-2023 | 0.238^**^ | 0.417^***^ | 0.17 | 0.422^***^ | 0.238^***^ | 0.434^***^ | 0.257^**^ | 0.482^***^ |
|  | [0.072,0.404] | [0.256,0.578] | [-0.011,0.351] | [0.260,0.585] | [0.087,0.388] | [0.286,0.581] | [0.101,0.413] | [0.323,0.641] |
| Variable | 0.144 | 0.062 | 0.14 | 0.096 | 0.091 | 0.274 | 0.052 | -0.387 |
|  | [-0.011,0.299] | [-0.085,0.210] | [-0.010,0.290] | [-0.056,0.249] | [-0.366,0.549] | [-0.117,0.666] | [-0.499,0.603] | [-1.157,0.384] |
| Variable * Period 2009-2013 | -0.151 | -0.14 | -0.047 | -0.118 | -0.238 | 0.629 | 0.457 | 0.064 |
|  | [-0.360,0.058] | [-0.322,0.042] | [-0.268,0.173] | [-0.343,0.108] | [-0.970,0.493] | [-0.287,1.546] | [-0.006,0.920] | [-0.255,0.384] |
| Variable * Period 2019-2023 | 0.077 | 0.051 | 0.173 | -0.064 | 0.164 | -0.774 | -0.113 | -0.349 |
|  | [-0.151,0.306] | [-0.188,0.291] | [-0.052,0.399] | [-0.312,0.184] | [-0.402,0.732] | [-2.863,1.314] | [-0.567,0.342] | [-0.749,0.052] |
| N | 6371 | 6940 | 5385 | 5346 | 4897 | 4925 | 4897 | 4925 |
|  | **Depression** | | **Diabetes** | | **Epilepsy** | | **High blood pressure** | |
|  | Female | Male | Female | Male | Female | Male | Female | Male |
| Period 2009-2013 | -0.135 | 0.030 | -0.158 | 0.068 | -0.141 | 0.053 | -0.153 | 0.055 |
|  | [-0.294,0.024] | [-0.093,0.153] | [-0.324,0.008] | [-0.069,0.205] | [-0.305,0.023] | [-0.084,0.190] | [-0.318,0.012] | [-0.080,0.191] |
| Period 2019-2023 | 0.273^**^ | 0.457^***^ | 0.237^**^ | 0.430^***^ | 0.247^**^ | 0.424^***^ | 0.250^***^ | 0.429^***^ |
|  | [0.124,0.422] | [0.312,0.603] | [0.089,0.385] | [0.283,0.578] | [0.099,0.395] | [0.276,0.572] | [0.102,0.397] | [0.281,0.577] |
| Variable | 0.350^*^ | 0.278 | 0.098 | -0.796 | 1.095^*^ | -12.164^***^ | 0.067 | -0.356 |
|  | [0.567,0.643] | [-0.005,0.563] | [-0.643,0.839] | [-2.379,0.787] | [0.200,1.991] | [-14.902,-9.425] | [-0.533,0.668] | [-1.339,0.627] |
| Variable * Period 2009-2013 | -0.397 | 0.028 | -0.016 | -1.148 | -0.77 | 12.915^***^ | -0.23 | 0.469 |
|  | [-0.519,0.440] | [-0.319,0.375] | [-0.727,0.695] | [-2.674,0.378] | [-1.733,0.193] | [11.652,14.177] | [-1.139,0.680] | [-1.306,2.244] |
| Variable * Period 2019-2023 | -0.003 | -0.546 | 0.371 | -0.082 | -0.987 | 13.064^***^ | -14.572^***^ | 0 |
|  | [-0.375,0.367] | [-1.162,0.70] | [-0.781,1.523] | [-1.641,1.477] | [-2.317,0.343] | [11.603,14.525] | [-16.041,-13.104] | [0.000,0.000] |
| N | 4897 | 4925 | 4897 | 4925 | 4897 | 4925 | 4897 | 4925 |
|  | **Age group** | | **Child responsibility** | | **Incomes** | | **IMD** | |
|  | Female | Male | Female | Male | Female | Male | Female | Male |
| Period 2009-2013 | 0.037 | 0.144^**^ | -0.204^**^ | 0.033 | -0.175^*^ | 0.039 | -0.138 | -0.042 |
|  | [-0.095,0.168] | [0.035,0.254] | [-0.339,-0.069] | [-0.077,0.143] | [-0.311,-0.039] | [-0.077,0.155] | [-0.294,0.017] | [-0.165,0.080] |
| Period 2019-2023 | 0.176^*^ | 0.276^***^ | 0.260^***^ | 0.423^***^ | 0.214^*^ | 0.381^***^ | 0.196^*^ | 0.403^***^ |
|  | [0.017,0.335] | [0.124,0.428] | [0.108,0.412] | [0.281,0.564] | [0.048,0.380] | [0.224,0.539] | [0.010,0.382] | [0.227,0.579] |
| Variable | -0.196^*^ | -0.297^***^ | -0.291^*^ | 0.355 | -0.086 | 0.097 | 0.08 | -0.094 |
|  | [-0.385,-0.007] | [-0.469,-0.125] | [-0.555,-0.026] | [-0.147,0.857] | [-0.220,0.048] | [-0.026,0.221] | [-0.180,0.341] | [-0.382,0.194] |
| Variable * Period 2009-2013 | -0.241 | -0.068 | 0.281 | 1.722 | 0.116 | -0.108 | -0.005 | 0.114 |
|  | [-0.494,0.011] | [-0.291,0.156] | [-0.042,0.603] | [-0.293,3.737] | [-0.081,0.313] | [-0.275,0.059] | [-0.233,0.222] | [-0.087,0.315] |
| Variable * Period 2019-2023 | 0.156 | 0.553^***^ | 0.059 | -1.066 | 0.145 | 0.135 | 0.209 | 0.092 |
|  | [-0.172,0.484] | [0.243,0.863] | [-0.369,0.488] | [-3.127,0.995] | [-0.072,0.362] | [-0.086,0.355] | [-0.057,0.475] | [-0.179,0.364] |
| N | 6403 | 6963 | 5881 | 6026 | 6196 | 6797 | 6399 | 6954 |

### Supplementary file S12. Fixed effect Poisson Regression by age group (16-19, 20-24)

|  | Health | | Psychological distress | | Multimorbidity | | Asthma | |
| --- | --- | --- | --- | --- | --- | --- | --- | --- |
|  | 16-19 | 20-24 | 16-19 | 20-24 | 16-19 | 20-24 | 16-19 | 20-24 |
| Period 2009-2013 | -0.390^***^ | 0.197^**^ | -0.305^*^ | 0.237^***^ | -0.448^***^ | 0.195^**^ | -0.452^**^ | 0.195^*^ |
|  | [-0.592,-0.188] | [0.077,0.318] | [-0.565,-0.045] | [0.100,0.375] | [-0.708,-0.188] | [0.047,0.344] | [-0.732,-0.171] | [0.022,0.367] |
| Period 2019-2023 | 0.452^***^ | 0.126 | 0.371^**^ | 0.124 | 0.434^***^ | 0.107 | 0.473^***^ | 0.112 |
|  | [0.194,0.710] | [-0.030,0.282] | [0.092,0.651] | [-0.033,0.281] | [0.185,0.682] | [-0.034,0.249] | [0.212,0.734] | [-0.034,0.259] |
| Variable | 0.01 | -0.017 | 0.106 | 0.046 | 0.241 | 0.195 | 0.179 | -0.183 |
|  | [-0.239,0.260] | [-0.146,0.112] | [-0.145,0.357] | [-0.085,0.176] | [-0.675,1.157] | [0.136,0.527] | [-0.875,1.233] | [-0.823,0.457] |
| Variable * Period 2009-2013 | 0.004 | -0.12 | -0.186 | -0.03 | NA | -0.259 | 0.021 | -0.036 |
|  | [-0.306,0.313] | [-0.286,0.046] | [-0.525,0.153] | [-0.231,0.170] |  | [-0.987,0.468] | [-0.643,0.685] | [-0.368,0.295] |
| Variable * Period 2019-2023 | 0.122 | -0.007 | 0.203 | 0.042 | 0.554 | -0.439 | -0.205 | -0.124 |
|  | [-0.263,0.507] | [-0.203,0.189] | [-0.159,0.565] | [-0.161,0.245] | [-0.633,1.743] | [-1.092,0.212] | [-0.928,0.519] | [-0.616,0.367] |
| N | 3244 | 6538 | 2565 | 5280 | 3260 | 4640 | 2518 | 4640 |
|  | Depression | | Diabetes | | Epilepsy | | High blood pressure | |
|  | 16-19 | 20-24 | 16-19 | 20-24 | 16-19 | 20-24 | 16-19 | 20-24 |
| Period 2009-2013 | -0.358^***^ | 0.162^*^ | -0.462^***^ | 0.197^*^ | -0.450^***^ | 0.201^**^ | -0.456^***^ | 0.204^**^ |
|  | [-0.578,-0.138] | [0.024,0.300] | [-0.723,-0.200] | [0.046,0.347] | [-0.710,-0.190] | [0.053,0.350] | [-0.716,-0.196] | [0.056,0.352] |
| Period 2019-2023 | 0.491^***^ | 0.132 | 0.443^***^ | 0.099 | 0.433^***^ | 0.1 | 0.443^***^ | 0.102 |
|  | [0.233,0.794] | [-0.008,0.273] | [0.197,0.689] | [-0.042,0.240] | [0.187,0.680] | [-0.042,0.241] | [0.197,0.689] | [-0.040,0.243] |
| Variable | 0.098 | 0.184 | 0 | 0.076 | 0.152 | 0.896 | -11.986^***^ | -0.096 |
|  | [-0.299,0.496] | [-0.040,0.408] | [0.000,0.000] | [-0.693,0.845] | [-0.344,0.648] | [-0.635,2.428] | [-13.946,-10.026] | [-0.627,0.435] |
| Variable * Period 2009-2013 | -0.212 | -0.096 | 12.791^***^ | -0.554 | 0.152 | -0.826 | 12.806^***^ | -0.793 |
|  | [-0.892,0.468] | [-0.465,0.273] | [10.813,14.768] | [-1.359,0.251] | [0.152,0.152] | [-1.953,0.302] | [10.837,14.775] | [-1.808,0.222] |
| Variable * Period 2019-2023 | -0.488 | -0.224 | 0 | 0.173 | 27.859^***^ | 0.378 | 0 | 0 |
|  | [-1.238,0.341] | [-0.556,0.108] | [0.000,0.000] | [-0.938,1.285] | [25.884,29.835] | [-2.165,2.920] | [0.000,0.000] | [0.000,0.000] |
| N | 2518 | 4640 | 2518 | 4640 | 2518 | 4640 | 2518 | 4640 |
|  | Child responsibility | | Incomes | | IMD | |  |  |
|  | 16-19 | 20-24 | 16-19 | 20-24 | 16-19 | 20-24 |  |  |
| Period 2009-2013 | -0.441^***^ | 0.236^***^ | -0.411^***^ | 0.187^**^ | -0.457^***^ | 0.163^*^ |  |  |
|  | [-0.661,-0.221] | [0.115,0.356] | [-0.616,-0.207] | [0.068,0.306] | [-0.691,-0.223] | [0.028,0.297] |  |  |
| Period 2019-2023 | 0.451^***^ | 0.116 | 0.452^***^ | 0.094 | 0.366^*^ | 0.119 |  |  |
|  | [0.205,0.696] | [-0.021,0.254] | [0.188,0.715] | [-0.059,0.247] | [0.080,0.653] | [-0.052,0.291] |  |  |
| Variable | -0.104 | 0.138 | -0.024 | -0.001 | -0.063 | 0.044 |  |  |
|  | [-0.684,0.476] | [-0.159,0.436] | [-0.224,0.175] | [-0.112,0.110] | [-0.573,0.447] | [-0.197,0.285] |  |  |
| Variable * Period 2009-2013 | -0.499 | -0.08 | 0.068 | -0.051 | 0.171 | 0.018 |  |  |
|  | [-1.648,0.651] | [-0.439,0.279] | [-0.212,0.348] | [-0.212,0.110] | [-0.184,0.526] | [-0.187,0.223] |  |  |
| Variable * Period 2019-2023 | -0.609 | -0.286 | 0.095 | 0.055 | 0.379 | 0.009 |  |  |
|  | [-1.875,0.657] | [-0.717,0.144] | [-0.257,0.448] | [-0.130,0.240] | [-0.144,0.902] | [-0.242,0.260] |  |  |
| N | 2900 | 5840 | 3193 | 6396 | 3257 | 6569 |  |  |
